# Genomic epidemiology reveals post-vaccine re-emergence of vaccine-associated G1P[8] rotavirus lineages with variable disease severity

**DOI:** 10.64898/2026.09.24.26362359

**Authors:** Chimwemwe Mhango, End Chinyama, Akuzike Banda, Richard Wachepa, Landilani Gauti, Ernest Matambo, Flywell Kawonga, Wanangwa Ndovie, David Chaima, Allan Zuza, Akuzike Kalizang’oma, Mwiza Mhango, Nigel A. Cunliffe, Nkonga Joyce Tshiswaka, Milton T Mogotsi, Ayodeji E Ogunbayo, Francis E. Dennis, Martin M. Nyaga, Benjamin Kumwenda, Celeste M. Donato, Arox W. Kamng’ona, Chrispin Chaguza, Khuzwayo C. Jere

## Abstract

Group A rotavirus G1P[8] strains, which are targeted by the Rotarix vaccine, have persisted in several countries following the introduction of the Rotarix vaccine. However, the evolutionary origins of the persisting strains remain unclear. Here, we leveraged over two decades of rotavirus surveillance data from Malawi to understand the genomic epidemiology of G1P[8] strains in a high-disease-burden sub-Saharan African setting. We observed a reversion of G1P[8] strains to the conventional Wa-like genotype constellation identified pre-vaccination, following transient circulation of DS-1-like G1P[8] strains. Most G1P[8] lineages that re-emerged post-vaccination circulated before vaccine introduction. Furthermore, new strains emerged post-vaccination through re-assortment of pre-existing viruses. The predominant post-vaccination lineage (MW-VP7-L2) contained an N147D substitution at an antigenically important site of the VP7 glycoprotein and showed evidence of episodic diversifying selection. Children infected with the MW-VP7-L2 lineage had a higher severity score of gastroenteritis than infections with other co-circulating lineages. These findings highlight the importance of long-term genomic surveillance to understand rotavirus transmission, evolution, and disease severity, and to guide public health interventions.

## Introduction

*Rotavirus alphagastroenteritidis,* or group A rotavirus (RVA), remains a leading cause of acute gastroenteritis among children under five years of age worldwide, accounting for approximately 128,500 deaths annually, particularly in low-resource settings^1^. The introduction of live-attenuated oral rotavirus vaccines has significantly reduced rotavirus deaths in under-five-year-olds globally^2,3^. While post-licensure studies have demonstrated high vaccine effectiveness (80–90%) in high-income countries (HICs)^4,5^, substantially lower estimates (45–60%) have been observed in low- and middle-income countries (LMICs)^4,5^. Several factors have been proposed to explain the reduced vaccine effectiveness in these settings, including high force of infection, vaccine neutralisation by maternal antibodies^6^, coinfections^7^, gut microbiome configuration^8^, malnutrition, poor sanitation and hygiene interventions^9^, environmental enteric dysfunction^10,11^, and the extensive genetic and antigenic diversity of rotaviruses resulting in poor heterotypic protection^12^.

RVA, a member of the *Sedoreoviridae* family, contains an eleven-segmented double-stranded RNA (dsRNA) genome^13^. The lack of proofreading during replication, due to its error-prone RNA-dependent RNA polymerase (encoded by the VP1 genome segment), facilitates the accumulation of point mutations, resulting in their extensive genomic diversity^14^. Besides point mutations, RVAs also undergo reassortment of their genomic segments upon co-infection of the same host cell with different strains, producing new constellations of genotypes associated with the segments^14,15^. Although less common, intragenic recombination through template switching can also occur during RVA replication, generating mosaic genome segments that similarly increase viral diversity in the population^14^. In addition, RVAs can infect a wide range of hosts^13^, creating opportunities for zoonotic and reverse zoonotic transmission^14,16,17^, which introduce novel RVA genomic segments into human populations. Collectively, these evolutionary mechanisms underpin RVA’s high genetic diversity. Whilst RVA genomic diversity is relatively straightforward to characterize at broader levels (i.e., the genotype), variation at the sub-genotype level (i.e., the lineage) remains poorly defined, partly due to the lack of universal RVA lineage nomenclature, which complicates global genomic surveillance and epidemiology.

Several viral pathogens have been extensively studied to directly link genomic variation with clinical and epidemiological outcomes. For example, large-scale genomic surveillance of the severe acute respiratory syndrome coronavirus 2 (SARS-CoV-2) during the pandemic enabled the rapid identification and tracking of variants of concern, including the Delta and Omicron variants, which were associated with increased transmissibility^18,19^, altered disease severity^20–22^, and substantial immune escape^23–25^. These evolutionary changes directly affected SARS-CoV-2 vaccine development and effectiveness^26,27^, necessitating regular updates to vaccine formulations and booster strategies^28^. Similarly, differences in disease severity have been reported among clades of the 2009 pandemic influenza A(H1N1) virus across different study settings^29^. Notably, the emergence of oseltamivir-resistant H1N1 influenza strains^30,31^ and molnupiravir-treatment-associated SARS-CoV-2 strains^32^ demonstrates how gradual viral evolution can directly compromise therapeutic effectiveness, underscoring the importance of integrating genomic surveillance into clinical and public health. Despite the growing number of RVA genomic studies globally, none has systematically assessed changes in RVA vaccine-induced population structure at the lineage level and evaluated the impact of sub-genotypic or lineage-specific differences on clinical outcomes and vaccine performance.

Despite RVA vaccine use for over two decades, RVA-associated gastroenteritis still persists worldwide^33^. Understanding how RVA evolution influences disease outcomes and vaccine effectiveness therefore remains a critical challenge, particularly in the LMIC settings where the disease burden remains high^28^. Integrating genomic, epidemiological, and clinical data provides a robust but underutilised framework for addressing this knowledge gap. Here, we leverage over 20 years of rotavirus surveillance data (1997-2019) from Malawi, a high disease-burden country in sub-Saharan Africa, spanning the periods before and after Rotarix RVA vaccine introduction, to investigate the genomic epidemiology and evolutionary dynamics of vaccine-associated G1P[8] strains. We further evaluate whether changes in RVA genomes and circulating lineages are associated with differences in the clinical severity of RVA-associated gastroenteritis in this setting.

## Results

### Leveraging over two decades of RVA surveillance data to monitor disease burden and strain dynamics in under-fives

We leveraged over two decades of sentinel hospital-based RVA surveillance in a high-disease-burden sub-Saharan African setting at the Queen Elizabeth Central Hospital (QECH), the largest tertiary referral hospital in Blantyre district, Malawi (**Figure 1a**). This setting offers a rare opportunity for population-level epidemiological studies and clinical interventions, as QECH serves as the main teaching hospital in southern Malawi and is the primary referral facility for 32 community-based health centres within the district^34^. We have conducted hospital-based RVA surveillance at QECH continuously since 1997, except for a temporary interruption between 2010 and 2011, when we conducted no surveillance (**Figure 1b**). To reduce the burden of RVA-associated gastroenteritis, the government of Malawi introduced a live attenuated G1P[8] Rotarix rotavirus vaccine (GlaxoSmithKline, Rixensart, Belgium) into its expanded immunisation programme on 28 October 2012. Consequently, we classified samples collected before and after this date as pre- and post-vaccine specimens, respectively.

**Figure 1.**
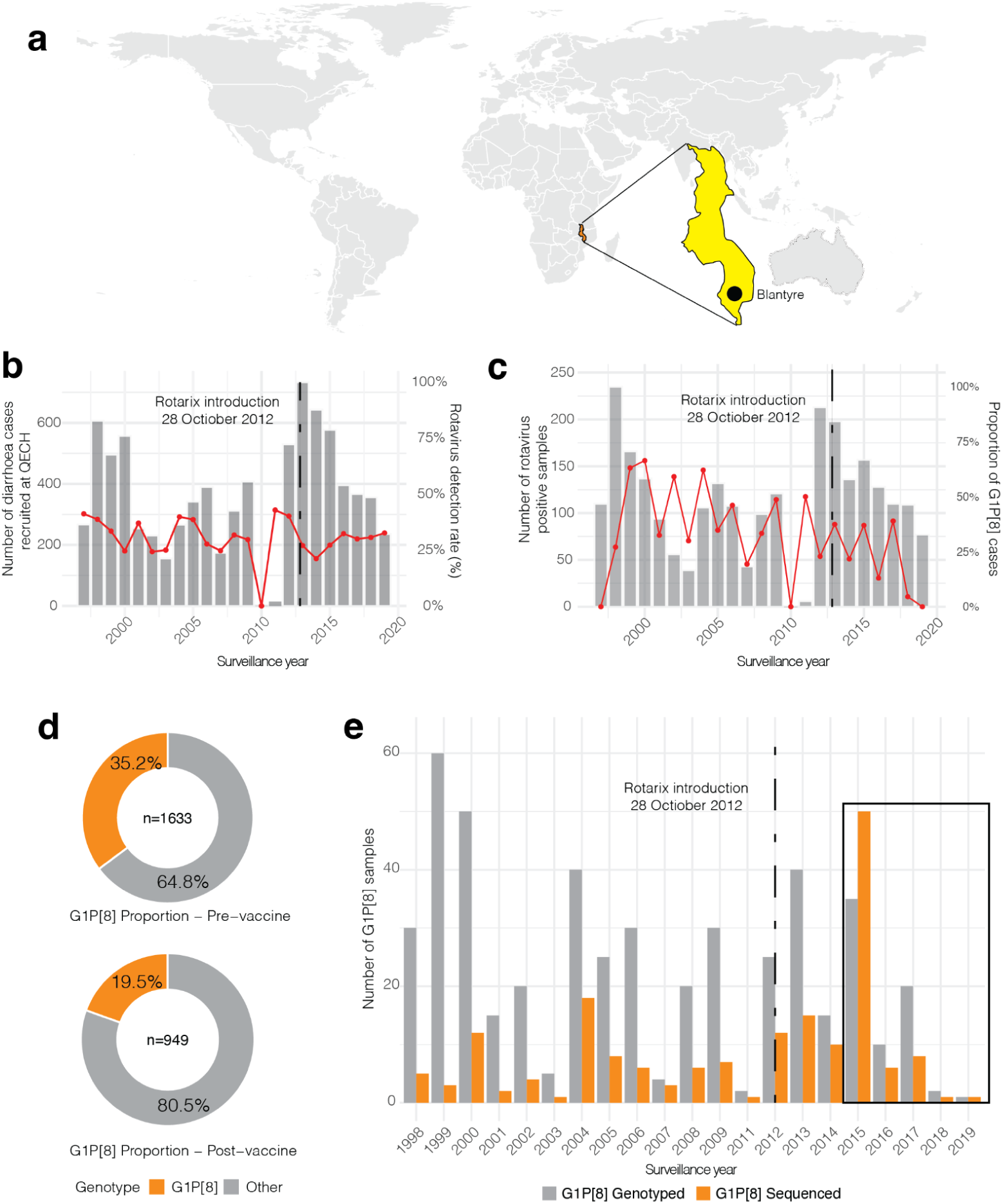
Rotavirus surveillance and temporal distribution of G1P[8] strains in Blantyre, Malawi, 1997–2019. **(a)** Geographic location of Blantyre in southern Malawi, where rotavirus surveillance was conducted at Queen Elizabeth Central Hospital (QECH). **(b)** Annual number of children with diarrhoea recruited at QECH (left y-axis) and the proportion that tested positive for rotavirus(right y-axis). **(c)** Annual number of rotavirus-positive samples (left y-axis) and the proportion of genotyped rotavirus cases identified as G1P[8] (right y-axis). **(d)** Proportion of genotyped rotavirus samples classified as G1P[8] or other genotypes during the pre-vaccine and post-vaccine period. **(e)** Annual number of G1P[8] samples successfully genotyped and the subset for which complete whole-genome sequences were generated.

We collected 8,322 stool specimens during the surveillance period (1997-2019), including 4,896 pre-vaccine samples and 3,426 post-vaccine samples. Enzyme immunoassays detected RVA in 33.4% (1,633 of 4,896) of pre-vaccine and 27.7% (949 of 3,426) of post-vaccine samples. Genotyping of RVA-positive samples based on the VP7 (designated G genotype) and VP4 (designated P genotype) outer capsid genes revealed substantial genotype diversity throughout the surveillance period^35^. This diversity resulted in multiple G and P genotype combinations, with marked temporal fluctuations in their relative frequencies^35^. Notably, G1P[8] strains, corresponding to the same G–P genotypes as the Rotarix vaccine strain, circulated consistently in Malawi from around 1998 until 2019, after which they were no longer detected in the surveillance population (**Figure 1c**). We identified 760 G1P[8] strains over the surveillance period, comprising 575 pre-vaccine and 185 post-vaccine strains (**Figure 1d**). This comprehensive longitudinal collection of G1P[8] strains spanning both the pre- and post-vaccine periods provided a unique population for investigating long-term evolutionary changes in this vaccine-targeted genotype in the context of Rotarix introduction.

We collated whole-genome sequencing datasets of G1P[8] strains collected before and after Rotarix vaccine introduction to investigate the long-term evolutionary dynamics of G1P[8] strains and the factors contributing to their continued circulation in Malawi. We obtained previously generated whole-genome sequences from G1P[8] strains collected between 1998 and 2014 from an earlier study^36^. To extend genomic surveillance into the later post-vaccine period, we generated new whole-genome sequences of 77 G1P[8] strains collected between 2015 and 2019 for in-depth downstream analyses (**Figure 1e and Table S1**). We combined these newly generated post-vaccine genomes with previously sequenced pre-vaccination strains to create a longitudinal genomic dataset comprising 215 whole genomes spanning over two decades of G1P[8] strain circulation in Malawi (**Table 1**). This extensive dataset allowed investigation of the evolution and persistence of G1P[8] strains across the pre- and post-vaccine periods.

**Table 1.**
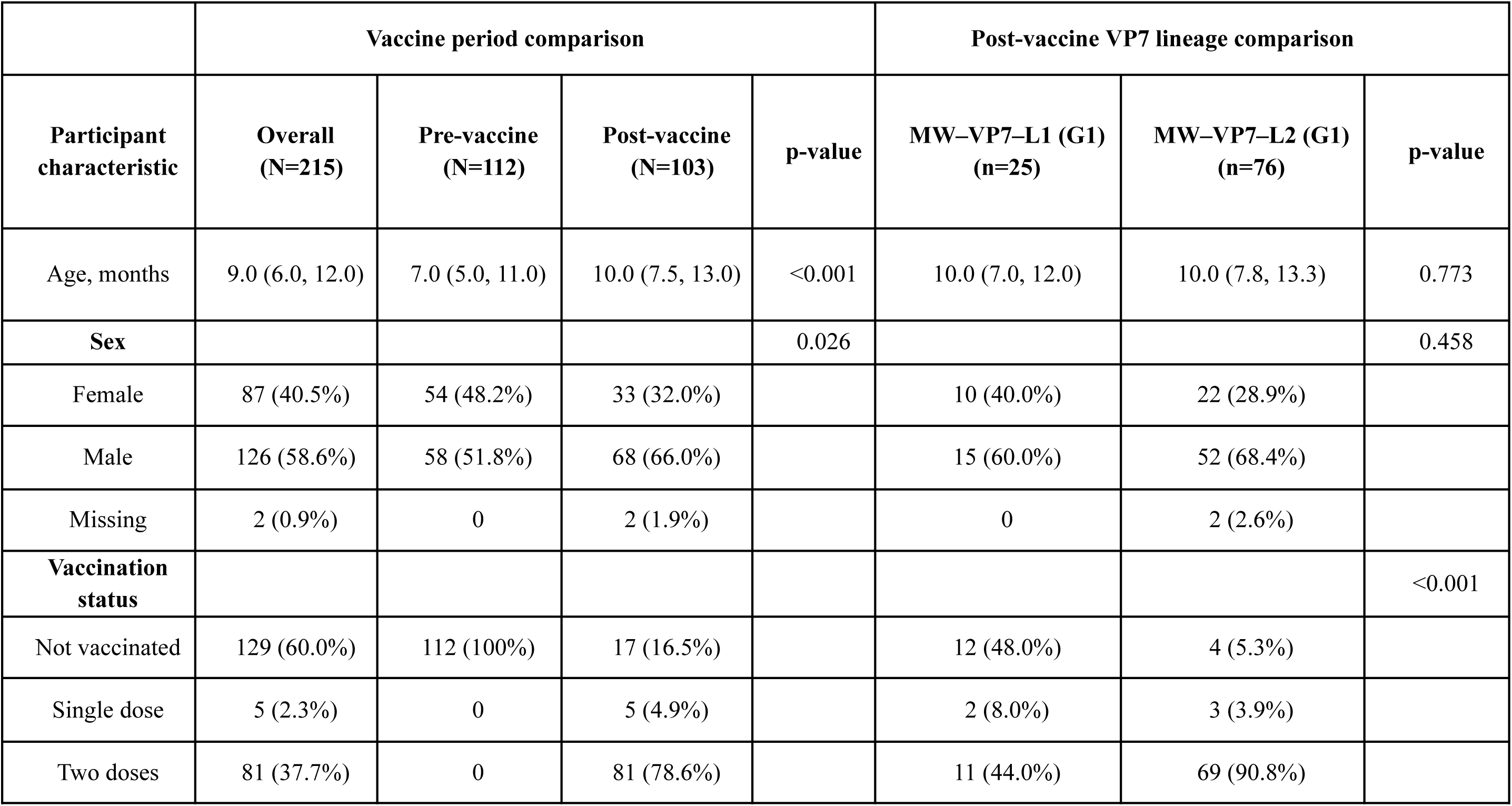
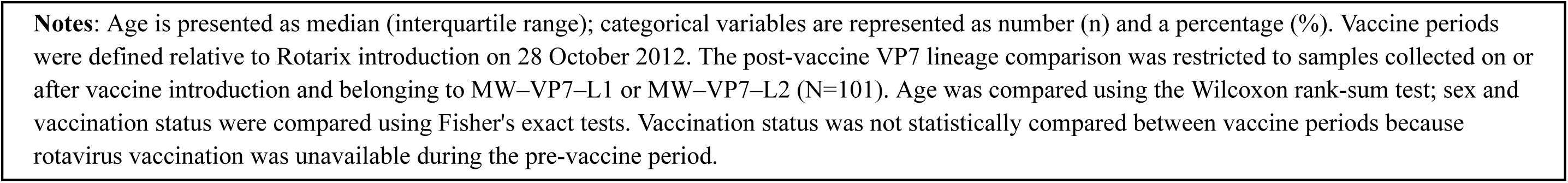
Participant characteristics by vaccine period and post-vaccine VP7 lineage.

|  | Vaccine period comparison |  |  |  | Post-vaccine VP7 lineage comparison |  |  |
| --- | --- | --- | --- | --- | --- | --- | --- |
| Participant characteristic | Overall (N=215) | Pre-vaccine (N=112) | Post-vaccine (N=103) | p-value | MW–VP7–L1 (G1) (n=25) | MW–VP7–L2 (G1) (n=76) | p-value |
| Age, months | 9.0 (6.0, 12.0) | 7.0 (5.0, 11.0) | 10.0 (7.5, 13.0) | <0.001 | 10.0 (7.0, 12.0) | 10.0 (7.8, 13.3) | 0.773 |
| **Sex** |  |  |  | 0.026 |  |  | 0.458 |
| Female | 87 (40.5%) | 54 (48.2%) | 33 (32.0%) |  | 10 (40.0%) | 22 (28.9%) |  |
| Male | 126 (58.6%) | 58 (51.8%) | 68 (66.0%) |  | 15 (60.0%) | 52 (68.4%) |  |
| Missing | 2 (0.9%) | 0 | 2 (1.9%) |  | 0 | 2 (2.6%) |  |
| **Vaccination status** |  |  |  |  |  |  | <0.001 |
| Not vaccinated | 129 (60.0%) | 112 (100%) | 17 (16.5%) |  | 12 (48.0%) | 4 (5.3%) |  |
| Single dose | 5 (2.3%) | 0 | 5 (4.9%) |  | 2 (8.0%) | 3 (3.9%) |  |
| Two doses | 81 (37.7%) | 0 | 81 (78.6%) |  | 11 (44.0%) | 69 (90.8%) |  |

**Notes:** Age is presented as median (interquartile range); categorical variables are represented as number (n) and a percentage (%). Vaccine periods were defined relative to Rotarix introduction on 28 October 2012. The post-vaccine VP7 lineage comparison was restricted to samples collected on or after vaccine introduction and belonging to MW-VP7-L1 or MW-VP7-L2 (N=101). Age was compared using the Wilcoxon rank-sum test; sex and vaccination status were compared using Fisher's exact tests. Vaccination status was not statistically compared between vaccine periods because rotavirus vaccination was unavailable during the pre-vaccine period.

### G1P[8] strains circulating more than three years after vaccine introduction predominantly possessed Wa-like constellations

We performed full-genome classification of RVA strains using the standardised genotype nomenclature system encompassing all 11 genome segments. Each segment was assigned an individual genotype to define the complete genotype constellation of each sequenced strain as Gx-P[x]-Ix-Rx-Cx-Mx-Ax-Nx-Tx-Ex-Hx, where Gx represents the VP7 segment, P[x] for the VP4, Ix for VP6, Rx for VP1, Cx for VP2, Mx for VP3, Ax for NSP1, Nx for NSP2, Tx for NSP3, Ex for NSP4, and Hx for NSP5 segments^37^. Although the VP7 (G-type) and VP4 (P-type) genomic segments of human RVAs encoding the outer capsid proteins exhibit extensive genomic diversity, the remaining segments generally segregate into two major backbone constellations: Wa-like (Gx-P[x]-I1-R1-C1-M1-A1-N1-T1-E1-H1; genogroup I) and DS-1-like (Gx-P[x]-I2-R2-C2-M2-A2-N2-T2-E2-H2; genogroup II)^38^. Our previous whole-genome analyses of Malawian RVA strains demonstrated that G1P[8] strains circulating before vaccine introduction (1998–2012) predominantly possessed the typical Wa-like genotype constellation, whereas G1P[8] strains detected shortly after Rotarix introduction (2013–2014) exhibited an atypical DS-1-like genotype constellation^36^. To determine whether these DS-1-like G1P[8] strains persisted beyond 2014, we characterised the complete genotype constellations of G1P[8] strains collected between 2015 and 2019. We successfully generated 77 complete genomes from strains collected during this period. Of these, 98.7% (76 out of 77) possessed the typical Wa-like genotype constellation [G1-P[8]-I1-R1-C1-M1-A1-N1-T1-E1-H1] (**Table S1**). The remaining strain, collected in 2015, also possessed a predominantly Wa-like genomic backbone (G1-P[8]-I1-R1-C1-M1-A2-N1-T1-E1-H1) but carried an A2 NSP1 genotype instead of the typical A1 genotype. As the A2 genotype is characteristically associated with DS-1-like strains, this constellation is consistent with a single-segment reassortment involving NSP1 (**Table S1**). Together, these findings show that the atypical DS-1-like G1P[8] strains observed shortly after introducing the Rotarix vaccine did not persist into the later post-vaccine period. Instead, G1P[8] strains circulating from 2015 onwards were almost exclusively characterised by the typical Wa-like genomic backbone that had predominated before vaccine introduction.

### Re-emerging post-vaccine Wa-like G1P[8] strains comprised both pre-existing and newly detected local lineages

Because G1P[8] strains detected more than three years after vaccine introduction predominantly possessed a Wa-like genotype constellation, we investigated their genetic relatedness at finer genomic resolution. We created a reference database comprising all available Malawian rotavirus genomes, including G1P[8] strains and other genotypes, and assigned local segment-specific lineages to each genome segment using a 98.5% nucleotide similarity threshold. Unlike genotype assignments based on the established whole-genome classification system^37^, which defines broad genetic categories, these local lineage assignments discriminate closely related variants within the same genotype. The high-resolution approach enables tracking of potentially persisting lineages, newly emerging variants, and strains arising from distinct introductions.

Across the entire surveillance period (1997-2019), the number of local lineages detected among G1P[8] strains ranged from three for NSP5 to eight for NSP3, indicating substantial variation in lineage diversity. Among strains sampled three years after vaccine introduction (2015-2019), we observed substantially lower lineage diversity, possibly due to the short sampling period, with individual genome segments containing between one and three local lineages (**Table S2**). The genomic segments associated with structural proteins (VP1, VP2, VP3, VP4, and VP7) each contained two lineages, whereas VP6 was represented by a single lineage. Among the non-structural protein genes, NSP4 was represented by one lineage; NSP2, NSP3, and NSP5 each contained two lineages, while NSP1 exhibited the greatest diversity with three distinct lineages (**Table S2**). These findings showed circulation of multiple local lineages at the sub-genotype level, reflecting repeated and continuous emergence of G1P[8] strains potentially through reemergence of rarely detected pre-existing lineages and genomic reassortment with locally circulating non-G1P[8] strains.

To determine the origins of this diversity, we next investigated whether individual segment-specific lineages represented persistence of lineages circulating before vaccine introduction and emergence lineages first detected during the post-vaccine period. Although we detected multiple lineages for most segments, a single lineage generally predominated during the post-vaccination period (2015-2019). We detected the predominant lineages associated with NSP2, NSP5, VP1, and VP2 genomic segments both before and after vaccine introduction, consistent with their continued circulation across the vaccine-introduction period. In contrast, the predominant lineages for NSP1, NSP3, VP3, VP4, VP6, and VP7 were first detected around the time of vaccine introduction and subsequently became established in the post-vaccine population. Notably, the predominant NSP4 lineage represented the re-emergence of a lineage last detected during the pre-vaccination period in 2006 (**Figure 2 and S1**). The other lineages occurred sporadically and generally disappeared within a year (**Figure 2 and S1**). Sporadically detected lineages associated with the NSP2, NSP3, and VP7 genomic segments had previously circulated in Malawi before vaccine introduction and were associated with Wa-like G1P[8] strains. In contrast, sporadic lineages associated with NSP1, NSP5, VP1, VP2, VP3, and VP4 were first detected three to four years after vaccine introduction (**Figure 2 and S1**). Thus, the segment-specific lineages associated with G1P[8] strains observed among later post-vaccine G1P[8] strains comprised a mixture of pre-existing pre-vaccine lineages and previously undetected lineages in Malawi.

**Figure 2.**
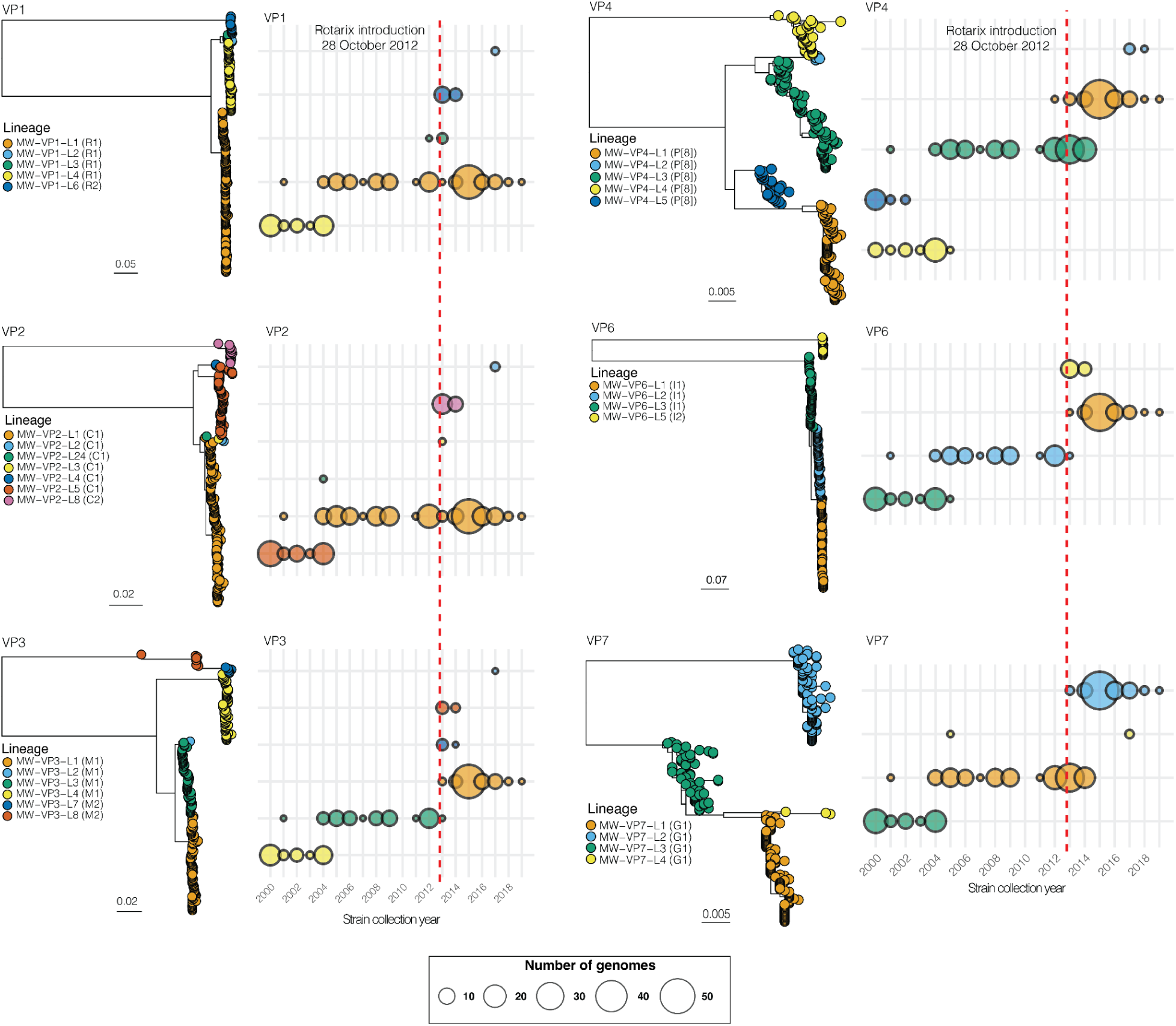
Classification and temporal distribution of local G1P[8] structural-protein gene lineages in Malawi, 1998–2019. Maximum-likelihood phylogenies are shown for VP1, VP2, VP3, VP4, VP6 and VP7. We assigned local Malawian lineages using ViralLC based on nucleotide sequence similarity across viruses sampled between 1998 and 2019; these designations represent locally defined lineages rather than global lineage nomenclature. Phylogenetic tips are coloured by assigned local lineage, with the corresponding conventional segment genotype shown in parentheses. For each structural-protein genome segment, sequences assigned to the same local lineage are generally clustered together in the maximum-likelihood tree, demonstrating concordance between the sequence-similarity-based classification and the inferred phylogenetic structure. The adjacent bubble plots show the annual distribution of each lineage, with bubble size proportional to the number of genomes detected in each surveillance year. The vertical dashed red line marks the introduction of the Rotarix rotavirus vaccine into Malawi’s national immunisation program on 28 October 2012. Phylogenetic scale bars indicate nucleotide substitutions per site.

Collectively, these findings demonstrate that the re-emergence of Wa-like G1P[8] strains following vaccine introduction did not result from the expansion of a single pre-vaccine viral lineage. Rather, the predominant post-vaccine G1P[8] population comprised a mosaic of segment-specific lineages with distinct temporal histories, including lineages that persisted across the vaccine-introduction period, lineages that emerged around the time of vaccine introduction and subsequently became established, or those that re-emerged post-vaccination after prolonged periods of cryptic circulation in the population.

### Predominant Wa-like G1P[8] lineages had pre-vaccine ancestry, suggesting cryptic circulation and subsequent introductions

Next, we estimated nucleotide substitution rates and the time to the most recent common ancestors (tMRCA) of the predominant local segment-specific lineages associated with post-vaccine Wa-like G1P[8] strains. This enabled us to estimate when these lineages began to diversify, approximate their minimum age, and to determine whether their emergence coincided with vaccine introduction. We observed substantial variation in the evolutionary rates across genome segments, ranging from 2.21×10^-4^ to 1.75×10^-3^ nucleotide substitutions per genomic site per year (**Figure 3a and S2**). This variation may reflect segment-specific differences in evolutionary constraints and selection pressures, although we cannot rule out potential biases from unequal sample sizes among predominant lineages. We observed the highest nucleotide substitution rates in the segments encoding structural proteins VP3, VP6, and VP7, whereas NSP5 showed the lowest rates (**Figure 3a and S2**), suggesting greater genetic stability compared with the more rapidly evolving segments.

**Figure 3.**
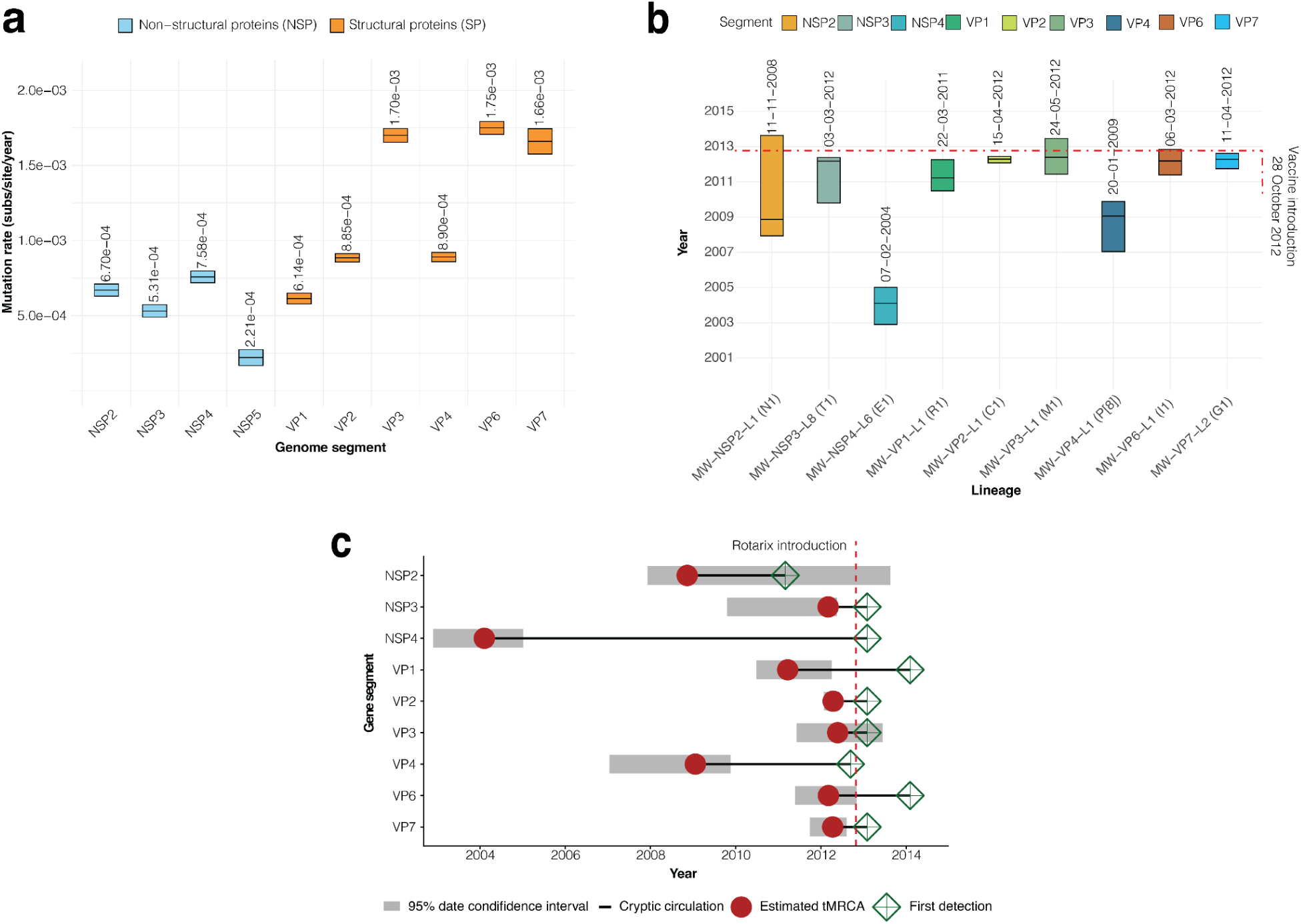
Evolutionary rates and origins of predominant post-2014 G1P[8] genome-segment lineages in Malawi. **(A)** Estimated nucleotide substitution rates for genome segments encoding non-structural proteins (NSP2–NSP5) and structural proteins (VP1–VP4, VP6 and VP7). Point estimates are shown for each segment, with uncertainty intervals representing the 95% confidence intervals. Rates are expressed as nucleotide substitutions per site per year. **(B)** Estimated times to the most recent common ancestor (tMRCAs) of the predominant G1P[8] local lineages detected after 2014. Boxes represent the estimated 95% date confidence intervals, and the horizontal line within each box indicates the tMRCA estimate. The red dashed horizontal line marks the time the Rotarix vaccine was introduced in Malawi on 28 October 2012. **(C)** Comparison of the estimated tMRCA and first observed detection date for each predominant post-2014 lineage. Red circles indicate the tMRCA estimates, grey bars show the corresponding 95% date confidence intervals, and open green diamonds indicate the first detection of each lineage in the surveillance dataset. Black horizontal lines between the upper confidence limit and the first detection date represent the potential period of cryptic circulation. The vertical dashed red line marks Rotarix introduction on 28 October 2012.

Using these evolutionary rates to estimate the tMRCA of the predominant Wa-like G1P[8] lineages revealed ancestral divergence times that largely predated Rotarix vaccine introduction, ranging from approximately 2008 to 2012, with most falling immediately before vaccine introduction (2011-2012) (**Figure 3b and S3**). The tMRCAs of the predominant lineages associated with NSP3, VP2, VP3, VP6, and VP7 genomic segments date to around 2012, close to but preceding vaccine introduction, while the VP1 lineage dates to approximately 2011 and the NSP2 and VP4 lineages to approximately 2009. In contrast, the predominant NSP4 lineage had a substantially older estimated tMRCA of approximately 2004 (**Figure 3b and S3**). We next compared the estimated tMRCAs of each predominant segment-specific lineage with the year the lineage was first detected through surveillance. For most segments, the inferred tMRCA preceded first detection by about 0.7 to 3.6 years, suggesting that these lineages circulated undetected for varying periods before surveillance captured them (**Figure 3c**). For example, the predominant NSP3, VP2, VP3, VP6, and VP7 lineages had tMRCAs dating to approximately 2012 but were not detected until 2013-2014. Similarly, the predominant VP1 and VP4 lineages remained undetected for approximately three years following their estimated ancestral divergence. The NSP4 lineage showed a considerably longer interval between its estimated tMRCA and subsequent detection, consistent with the historical circulation followed by prolonged non-detection and later re-emergence post-vaccination (**Figure 3c**).

Collectively, these findings demonstrate that the predominant segment-specific lineages associated with the post-vaccine Wa-like G1P[8] population had ancestral origins predating the introduction of the Rotarix vaccine. Their delayed detection is consistent with periods of cryptic circulation or incomplete sampling, while lineages first observed during the post-vaccine period may additionally reflect new introductions into Malawi. Thus, the later post-vaccine G1P[8] population appears to have risen from a combination of persistence and re-emergence of pre-existing viruses, along with potential introductions of previously unsampled lineages, rather than de novo emergence following vaccine introduction.

### Persistence of pre-existing Wa-like G1P[8] lineages at variable frequencies following post-vaccine introduction was associated with adaptive evolution of the VP7 glycoprotein

Next, we investigated whether the persistence of segment-specific Wa-like G1P[8] lineages in a highly vaccinated population (**Figure 4a**), with national rotavirus vaccine coverage approaching 90% ^39^, was associated with evidence of adaptive evolution. Given that the predominant post-vaccine Wa-like G1P[8] lineages largely emerged through persistence or re-emergence of pre-existing lineages rather than exclusively through new introductions, we hypothesised that differences in evolutionary mechanisms, including vaccine-associated selection pressures, may have shaped the variability in the frequency of the co-circulating Wa-like G1P[8] lineages post-vaccination, with only a subset expanding and becoming established in the population. We first screened all eleven genome segments for evidence of gene-wide episodic diversifying selection using the branch-site unrestricted statistical test for episodic diversification (BUSTED)^40^. We estimated global nonsynonymous-to-synonymous substitution rate ratios (dN/dS, ω) below one across all segments of the Wa-like G1P[8] lineages, indicating purifying selection predominated across the Wa-like G1P[8] genome (**Figure 4b**). Nevertheless, we detected statistically significant evidence of episodic diversifying selection in NSP2, VP1, VP2, VP3, VP6, and VP7 genome segments, indicating that a subset of sites or branches experienced diversifying selection despite the overall dominance of purifying selection. Of the two outer capsid proteins that are major targets of neutralising antibodies, VP7 showed significant evidence of gene-wide episodic diversifying selection (BUSTED, *p* = 0.0066), whereas VP4 did not (**Figure 4b**). To identify individual codons contributing to the VP7 selection signal, we performed codon-level analysis using the Mixed Effects Model of Evolution (MEME)^41^. Although most VP7 codons showed no evidence of episodic diversifying selection, six codons showed significant or suggestive evidence of selection. Codons 147 and 148 showed statistically significant evidence (MEME, *p* = 0.0169 and *p* = 0.0145, respectively), while codons 28, 116, 233 and 247 showed weaker, non-statistically significant signals (**Figure 4c**). Because residues 147 and 148 lie within the major antigenic regions of the VP7 glycoprotein^42^, we next examined amino acid variation at these two sites across the four VP7 Wa-like lineages detected during the period. We hypothesised that lineage-specific changes at sites under episodic diversifying selection could potentially contribute to the differences in frequencies observed among persisting VP7 lineages following vaccine introduction. We compared amino acid profiles across four Wa-like VP7 lineages (MW–VP7–L1, MW–VP7–L2, MW–VP7–L3, and MW–VP7–L4), using the earliest sequenced Malawian G1P[8] strain (GenBank accession: MG181232.1) as the reference. We found that most amino acid substitutions occurred sporadically and at low frequency across the four lineages. The VP7:Q28R substitution was shared in all lineages, occurring in ∼87% of the MW–VP7–L2 lineage and in all sequences of other lineages. In contrast, the VP7:N147D substitution occurred almost exclusively within the predominant post-vaccine Wa-like VP7 lineage MW–VP7–L2 and was present in 97.5% of sequences in this lineage (**Figure 4d**).

**Figure 4.**
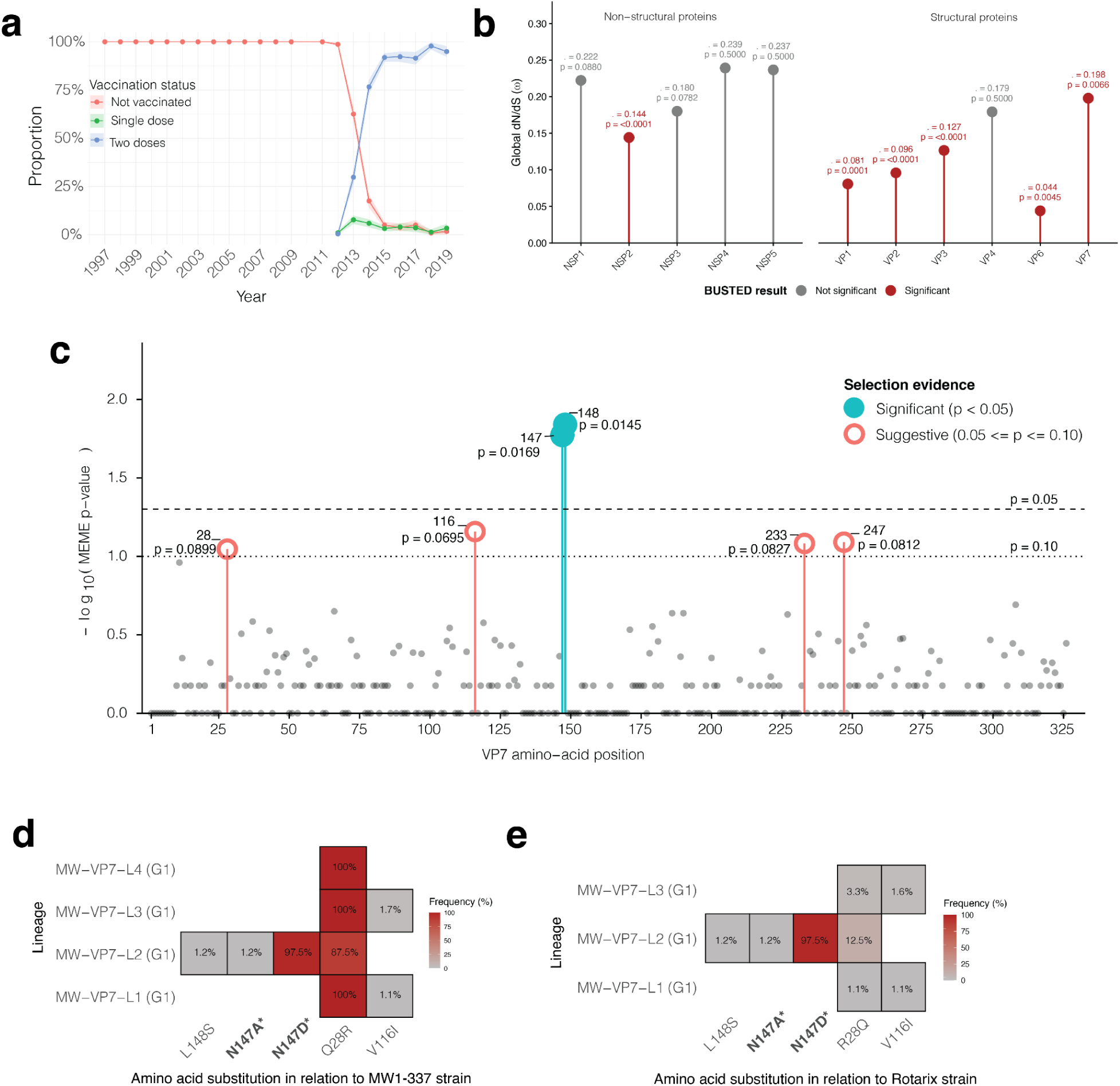
Evidence of adaptive evolution and lineage-associated amino acid variation among Malawian G1P[8] rotaviruses. **(a)** Annual Rotarix rotavirus vaccine coverage among children presenting with acute gastroenteritis at QECH from 1997 to 2019, shown as the proportion of unvaccinated or received one or two doses of Rotarix. **(b)** Gene-wide evidence of episodic diversifying selection in the outer-capsid proteins VP4 and VP7, assessed using BUSTED. VP7 showed significant evidence of selection (*p* = 0.0066), whereas VP4 was not significant (*p* = 0.50). The global nonsynonymous-to-synonymous rate ratios (ω) are shown above each protein. **(c)** Site-level evidence of episodic diversifying selection across the VP7 protein identified using MEME. Amino acid positions with significant evidence of selection (*p* < 0.05) are shown in cyan, while sites with suggestive evidence (0.05 ≤ *p* ≤ 0.10) are shown in salmon. The horizontal dashed lines indicate the *p* = 0.05 and *p* = 0.10 thresholds. **(d and e)** Heatmaps showing the frequency of selected VP7 amino acid substitutions within Malawian VP7 lineages. Values indicate the percentage of sequences within each lineage carrying the corresponding substitution, with darker shading representing a higher frequency. Asterisks identify substitutions occurring at sites detected by MEME as being under episodic diversifying selection

We next compared the amino acid profiles of the post-vaccine VP7 lineages associated with Wa-like backbone genes with the VP7 sequence of the G1P[8] Rotarix vaccine strain to determine whether the predominant post-vaccine Wa-like VP7 lineage differed from the vaccine strain at antigenic region positions that show episodic diversifying selection. We found that most amino acid substitutions were either shared among lineages or occurred at low frequencies, consistent with our findings comparing post-vaccine Wa-like lineages with the local pre-vaccine reference. Notably, however, the VP7:N147D substitution, present in ∼97.5% of the predominant post-vaccine Wa-like VP7 lineage (MW–VP7–L2), was absent in the Rotarix vaccine strain (**Figure 4e**).

Collectively, these findings show that the persistence and predominance of specific Wa-like G1P[8] lineages following vaccine introduction coincided with evidence of adaptive evolution in the VP7 gene. While these observations do not directly establish vaccine-driven selection, they suggest the possibility that vaccine-associated immune pressure may have contributed to the differential persistence and expansion of specific G1P[8] lineages in the post-vaccine population.

### The VP7:N147D substitution maps to an antigenic region of VP7 and increased in frequency following vaccine introduction in Malawi

Since the VP7:N147D mutation distinguished the predominant post-vaccine MW–VP7–L2 lineage from both the local pre-vaccine reference and the Rotarix vaccine strain, we next investigated its structural context and broader temporal distribution. We mapped the VP7 sites identified by MEME onto structural models of the Rotarix vaccine strain and representative post-vaccine MW–VP7–L1 and MW–VP7–L2 proteins (**Figure 5a and 5b**). The six sites showing significant or suggestive evidence of episodic diversifying selection were distributed across the VP7 structure, with residues 147 and 148 located within an exposed 7-2 antigenic region^43^ of the VP7 head domain. Comparison of the three structures showed that residue 147 differed between the predominant MW–VP7–L2 lineage and both Rotarix and MW–VP7–L1 lineage, with asparagine (VP7:N147) in Rotarix and MW–VP7–L1 lineage replaced by aspartic acid (VP7:D147) in MW–VP7–L2 lineage (**Figure 5b**). This placed the lineage-defining VP7:N147D substitution identified in our sequence analysis within a functionally relevant antigenic region of VP7.

**Figure 5.**
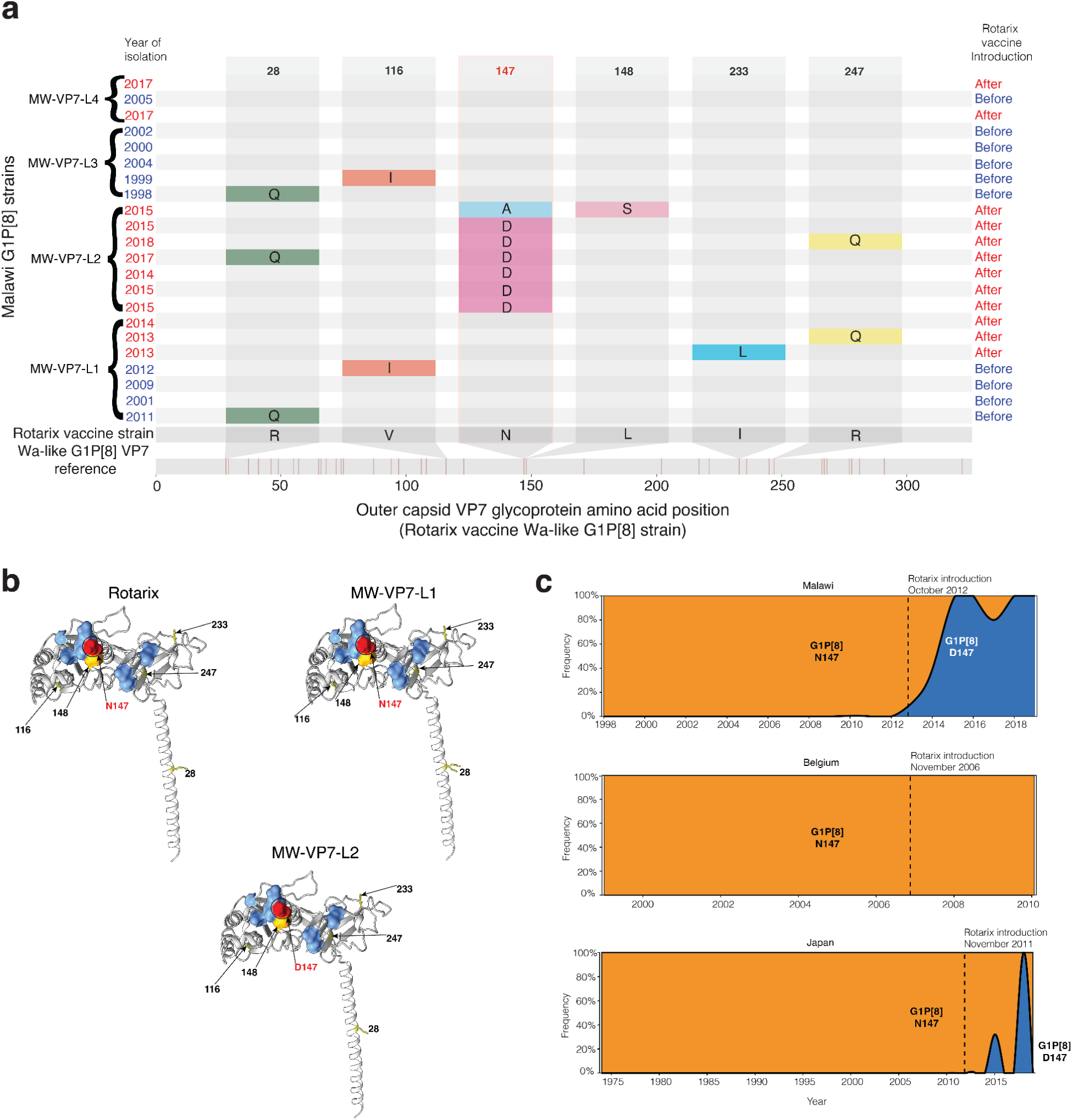
Lineage-specific amino-acid variation and structural localisation of sites under episodic diversifying selection in the VP7 glycoprotein of Malawian G1P[8] rotaviruses, relative to the VP7 of the Rotarix vaccine strain. **(a)** Amino-acid variation at VP7 codons identified by mixed-effects model of evolution (MEME) analysis. Codons 147 and 148 showed significant evidence of episodic diversifying selection, whereas codons 28, 116, 233 and 247 showed weaker/suggestive evidence. Representative Malawian G1P[8] sequences are grouped by VP7 lineage (MW-VP7-L1 to MW-VP7-L4), with isolation years coloured by vaccine period (blue, pre-vaccine; red, post-vaccine). Residues differing from the Rotarix VP7 sequence are indicated within coloured cells; unmarked cells indicate identity with Rotarix. **(b)** Structural localisation of the MEME-identified sites on superimposed VP7 models representing Rotarix, MW-VP7-L1 and MW-VP7-L2. The surface-exposed 7-2 neutralising antigenic region (residues 143, 145-148, 190, 217, 221 and 264) is highlighted. 147 and 148 lie within this region, with D147 distinguishing MW-VP7-L2 from MW-VP7-L1 and Rotarix, whereas positions 28, 116, 233, and 247 are indicated at their respective structural locations. (**c)** Temporal frequencies of G1P[8] viruses carrying asparagine (N147) or aspartic acid (D147) at VP7 residue 147 in Malawi, Belgium, and Japan. Residue frequencies were calculated from available G1P[8] VP7 sequences by year of isolation, with vertical dashed lines indicating Rotarix vaccine introduction in each setting.

We next examined whether the D147 variant emerged only in Malawi or also occurred among G1P[8] strains circulating in other vaccinated populations. Analysis of publicly available G1P[8] VP7 sequences showed a marked temporal shift from VP7:N147 to the D147 variant in Malawi following Rotarix introduction in October 2012 (**Figure 5c**). The VP7:N147 variant predominated among Malawian G1P[8] strains before vaccine introduction, whereas the frequency of the VP7:D147 mutation increased rapidly thereafter and became the predominant residue among strains sampled during the later post-vaccine period. This temporal transition corresponded with the expansion of MW–VP7–L2, in which the VP7:D147 variant was nearly fixed. In comparison, the temporal patterns differed in other populations examined. For instance, G1P[8] strains in Belgium maintained the VP7:N147 residue even after vaccine introduction, whereas Japanese G1P[8] strains showed the VP7:D147 variant during the later sampling period (**Figure 5c**). Additional analysis of Mozambican sequences similarly identified VP7:D147-allele-containing G1P[8] strains (**Figure S5**). Thus, the VP7:D147 variant was not unique to Malawi, but its rapid increase and subsequent predominance there after vaccine introduction were particularly pronounced.

Collectively, these findings indicate that the differential persistence of Wa-like G1P[8] lineages following Rotarix introduction coincided with adaptive evolution of the VP7 glycoprotein. The predominant post-vaccine MW–VP7–L2 lineage was characterised by the near-fixation of VP7:N147D, a substitution at a VP7 site under episodic diversifying selection that lies within a major antigenic region and differs from the residue present in the Rotarix vaccine strain. The pronounced post-vaccine increase of the D147 variant in Malawi, together with its occurrence in G1P[8] strains from other populations, suggests that this site is evolutionarily variable rather than uniquely Malawian. Although these observations do not establish vaccine-driven selection, the temporal, evolutionary, and structural evidence collectively support the hypothesis that changes at VP7 residue 147 may have contributed to the differential fitness of G1P[8] lineages circulating in the post-vaccine population.

### Post-vaccine G1P[8] viral populations were associated with differences in clinical disease severity

We next investigated whether G1P[8] lineages circulating after introduction of Rotarix vaccine were associated with differences in clinical disease severity. We hypothesised that the predominant Wa-like lineages would be associated with greater disease severity than less prevalent lineages. We restricted this analysis to the strains sampled during the post-vaccine period because detailed clinical severity data were unavailable for pre-vaccine samples, precluding direct comparison of disease severity before and after vaccine introduction. We grouped the post-vaccine strains by VP7 lineage, as the VP7 glycoprotein is a major outer-capsid antigen targeted by neutralising antibodies^42,44^ and encodes the G1 antigen shared by the circulating strains and the Rotarix vaccine strain^45^. This grouping of strains was further supported by our observation of episodic diversifying selection in VP7, although it does not necessarily imply that genomic variation in the VP7 protein alone determines clinical disease severity.

We compared the Vesikari scores, a standardised measure of disease severity associated with acute gastroenteritis^46^, among children infected with the two principal VP7 lineages of the post-vaccine Wa-like G1P[8] populations, MW–VP7–L1 and MW–VP7–L2. Of the 101 children included in the analysis, 25 were infected with the MW–VP7–L1 lineage and 76 with the MW–VP7–L2 lineage. Age and sex distributions were comparable between the two groups (**Table 1**). In contrast, vaccination status differed markedly by lineage (*p<0.001*): most children infected with the MW–VP7–L2 lineage received two doses of Rotarix, whereas those infected with the MW–VP7–L1 lineage were predominantly unvaccinated (**Table 1**). Consistent with our hypothesis, children infected with the predominant post-vaccine Wa-like MW–VP7–L2 lineage experienced more severe disease than those infected with the less prevalent MW–VP7–L1 lineage (**Figure 6a**). The median Vesikari score was 15.0 (IQR: 13.0–15.5) among children infected with MW–VP7–L2 compared with 13.0 (IQR: 11.0–13.2) among those infected with the MW–VP7–L1 lineage (*p<0.001*). Given the substantial difference in vaccination status between the two groups, we fitted a multivariable linear regression model to account for potential confounding by vaccination status/number of vaccine doses, age, and sex. After adjusting for these variables, we found that children infected with the MW–VP7–L2 lineage remained independently associated with a mean Vesikari score 2.32 points higher than that of MW–VP7–L1, whereas age, sex, and receipt of two vaccine doses were not significantly associated with overall disease severity (**Figure 6b**). We next examined the individual clinical components contributing to the Vesikari score to determine which features accounted for the observed differences in severity. We noted that children infected with the post-vaccine Wa-like MW–VP7–L2 lineage passed more stools per day, had an extended duration of diarrhoea, and were more dehydrated compared with those infected with MW–VP7–L1, whereas the remaining clinical features showed no apparent differences between the two lineages **(Figure S6**).

**Figure 6.**
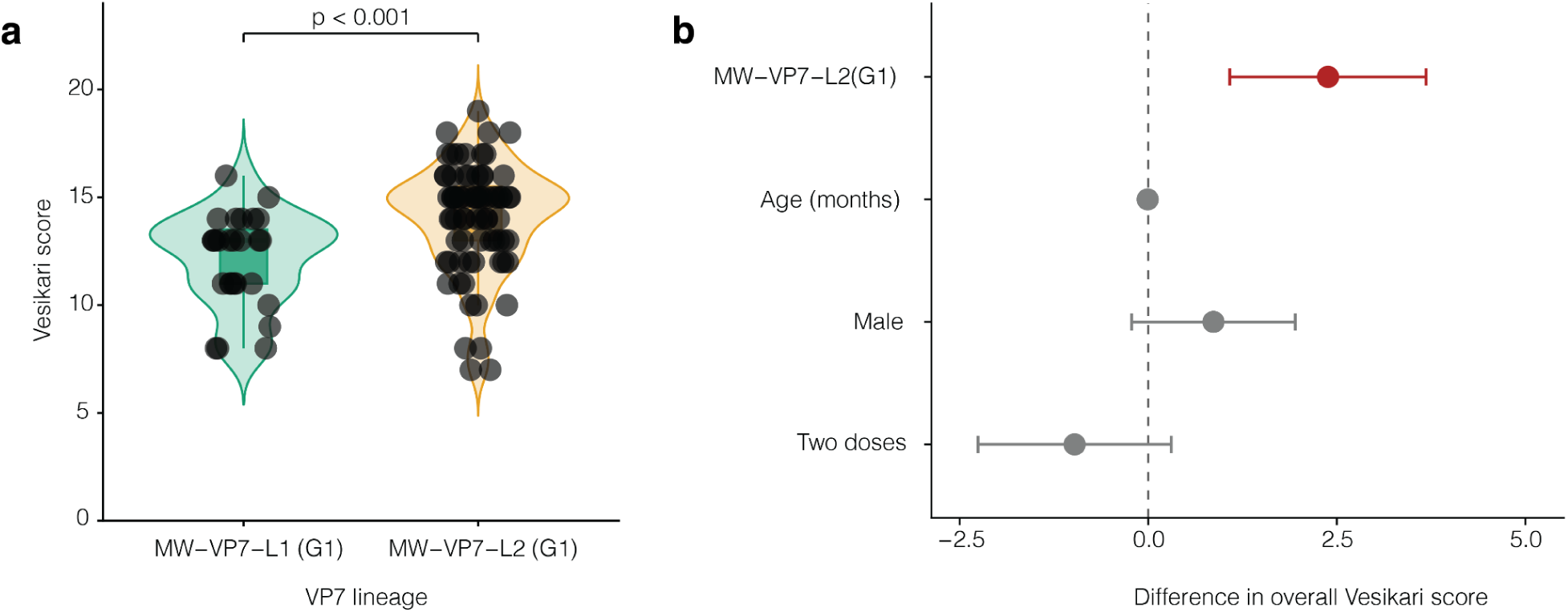
Association between G1P[8] viral lineage profile and rotavirus disease severity. **(a)** Distribution of overall Vesikari severity scores among children infected with viruses assigned to the MW–VP7–L1 (G1) and MW–VP7–L2 (G1) groups. We used the VP7 lineage designation to distinguish strains carrying the corresponding broader genomic lineage profiles. Violin plots show the score distributions with individual observations overlaid. **(b)** Multivariable linear regression estimates for differences in overall Vesikari score associated with viral lineage profile and participant covariates. Points represent regression coefficients and horizontal bars indicate 95% confidence intervals; the dashed vertical line denotes no difference. MW–VP7–L1 (G1), female sex, and fewer than two vaccine doses were reference categories, with age modelled continuously in months.

Together, these findings demonstrate that the two major post-vaccine Wa-like G1P[8] viral populations showed variable clinical disease severity, with the predominant MW–VP7–L2 lineage associated with more severe disease than other lineages. These observations highlight the value of integrating genomic surveillance with detailed clinical and epidemiological data to determine whether changes in viral populations are accompanied by clinically relevant phenotypic differences.

## Discussion

In this study, we leveraged over two decades of RVA genomic, epidemiological, and clinical surveillance to investigate the evolution and clinical characteristics of the G1P[8] strains circulating in Malawi after three years following introduction of the Wa-like G1P[8] Rotarix vaccine. Our study represents one of the longest longitudinal genomic investigations of a single rotavirus genotype within a single surveillance setting, providing a unique opportunity to reconstruct its evolutionary dynamics at whole-genome resolution across the pre- and post-vaccine periods. Several key findings emerged. First, G1P[8] strains circulating from three years after vaccine introduction predominantly possessed the conventional Wa-like genotype constellation that had circulated before vaccination, following the transient emergence of atypical DS-1-like G1P[8] strains shortly after vaccine introduction. Second, fine-scale, sub-genotypic, segment-level genomic analysis revealed distinct evolutionary histories of the post-vaccine Wa-like G1P[8] strains, with many lineages that later became established having originated or circulated, either detectably or cryptically, before vaccine introduction. Thus, post-vaccine Wa-like G1P[8] populations emerged largely through differential persistence, re-emergence, and reassortment of pre-existing RVA diversity rather than the emergence of an entirely novel constellation of genotypes. Third, the predominant post-vaccine Wa-like G1P[8] VP7 lineages carried a near-fixed amino acid substitution within a major antigenic epitope and at the site under episodic diversifying selection, raising the possibility that adaptive evolution contributed to its post-vaccine predominance. Finally, this predominant lineage was associated with greater clinical disease severity than a less prevalent co-circulating VP7 lineage. Together, these findings demonstrate the value of integrating longitudinal genomic surveillance with vaccination and clinical data to identify evolutionary changes in circulating viruses and determine their potential clinical consequences.

RVA G1P[8] strains have historically been among the most prevalent worldwide and were particularly dominant before the widespread introduction of the Rotarix vaccine^47,48^. Consistent with this global distribution, G1P[8] strains circulated in Malawi for over two decades, spanning both the pre- and post-vaccine periods^35,36^. Although their overall frequency declined following Rotarix introduction, G1P[8] strains continued to be detected in our long-term hospital-based surveillance seven years after vaccine introduction. The persistence and subsequent re-emergence of Wa-like G1P[8] strains are particularly notable because the Rotarix vaccine is derived from a live attenuated Wa-like G1P[8] strain that shares the same broad G and P genotypes as these circulating strains. Our early Rotarix vaccine effectiveness studies in Malawi estimated approximately 82% effectiveness against G1P[8] disease in a population in which national vaccine coverage exceeded 90% three years after vaccine introduction^39^. Continued circulation of Wa-like G1P[8] strains despite high vaccine coverage raised an important question of whether viruses classified as the same G1P[8] genotype remained genetically similar at finer, sub-genotypic resolution or represented distinct viral populations with different evolutionary histories.

Our findings show that continued detection of the re-emerging post-vaccine Wa-like G1P[8] strains largely reflected the persistence and reassembly of pre-existing viral diversity rather than replacement by an entirely new constellation of genotypes. Although these post-vaccine Wa-like G1P[8] strains appeared genetically similar under conventional genotype classification, detailed segment-level analysis revealed substantial sub-genotypic heterogeneity and distinct evolutionary histories across genome segments. Some lineages persisted across the pre- and post-vaccine periods, others appear to have circulated cryptically before becoming detectable, and reassortment brought together segments with different evolutionary histories within individual viruses. These dynamics resemble those observed for seasonal A influenza viruses, in which multiple lineages within the same subtype co-circulate, reassort, and undergo repeated replacement and reassortment over time^49–51^. These observations also align with our previous study of Malawian G1P[8] strains, which demonstrated the sequential replacement of distinct lineages in the pre-vaccine period and shortly after vaccine introduction^36^. The current analysis extends these observations by demonstrating continued lineage turnover during the later post-vaccine period and, importantly, that such turnover can occur independently across individual genome segments. These findings also highlight the limitations of conventional whole-genome genotype classification: viruses sharing an identical genotype constellation can harbour considerable sub-genotypic diversity that may be epidemiologically and biologically relevant. Standardised lineage nomenclatures at global and national levels could therefore complement existing genotype classification by enabling higher-resolution tracking of viral populations and linking genomic changes with epidemiological and clinical phenotypes.

A particularly notable finding was the predominance of the post-vaccine MW–VP7–L2 lineage and the near-fixation of the VP7:N147D substitution within the lineage. VP7, together with VP4, forms the outer capsid and is a major target of neutralising antibodies^52,43,53–55^. Amino acid changes within exposed VP7 epitopes could therefore affect recognition by vaccine-induced antibodies, making variations at these sites particularly relevant after introducing a G1P[8]-based vaccine. The VP7:N147D substitution lies within the VP7 7-2 neutralisation epitope^43^ and was present in almost all MW–VP7–L2 viruses but largely absent from the less prevalent post-vaccine Wa-like G1P[8] VP7 lineages. Moreover, codon 147 showed significant evidence of episodic diversifying selection. Experimental studies have previously implicated residue 147 in antibody-mediated neutralisation^42,56^, including evidence that Rotarix-induced monoclonal antibodies can neutralise viruses carrying certain residues at this position but fail to neutralise viruses carrying alternative amino acids^42,56^. However, these observations should not be interpreted as direct evidence that Rotarix selected for the VP7:N147D mutation. The post-vaccine success of MW–VP7–L2 could also reflect other viral characteristics, stochastic epidemiological processes, or selection acting elsewhere in the genome. Although our analyses found no evidence of episodic diversifying selection in VP4, we cannot entirely rule out the contributions from VP4 and other genome segments. Emerging evidence suggesting that VP7 may constitute a major target of human neutralising antibody responses and can independently induce neutralising antibodies^44^ nevertheless provides biological plausibility for the importance of VP7 variation. Importantly, the VP7:N147D-containing G1P[8] viruses have been reported in geographically diverse settings, including in South Africa^57^, Mozambique^58^, Japan, Thailand, and Vietnam^59^. The repeated observation of this substitution in post-vaccine populations warrants further investigation to determine whether it reflects convergent antigenic adaptation, the dissemination of related viral populations, or broader temporal changes in G1P[8] diversity. Together, our observations of the VP7:N147D mutation within a neutralising epitope, evidence of diversifying selection at this site, and its near fixation within the predominant post-vaccine lineage are consistent with the possibility that antigenic selection contributed to this lineage’s differential persistence post-vaccination.

A key strength of our study was integrating an extensive longitudinal genomic dataset with detailed epidemiological and clinical information, which allowed us to investigate whether genomic diversification within the post-vaccine Wa-like G1P[8] lineages was associated with differences in disease phenotype. Our previous studies from Malawi^60^ and elsewhere^61^ have reported differences in disease severity between broad RVA G- and P-genotype combinations, but whether or not viruses within the same genotype differ clinically remains poorly understood. Our findings show that children infected with strains carrying the predominant post-vaccine MW–VP7–L2 experienced more severe gastroenteritis than those infected with the less prevalent MW–VP7–L1, characterised particularly by more frequent and prolonged diarrhoea and greater dehydration. Importantly, the association remained after adjusting for age, sex, vaccination status, and number of vaccine doses. These findings suggest that genomic diversification within a single RVA genotype may be accompanied by clinically relevant phenotypic differences, as observed in genetically distinct viral populations of other pathogens, including influenza A(H1N1)pdm09 and SARS-CoV-2^29,62,63^. However, our analysis does not establish that the VP7:N147D mutation specifically caused the observed difference in severity. The VP7 lineage served as a marker of genetically distinct viral populations that may differ elsewhere in their genomes. A key consideration for future work will be conducting systematic viral genome-wide association studies (GWAS) of larger datasets to identify candidate genetic changes associated with viral characteristics, including disease severity, viral load, and effects of therapy, as done for SARS-CoV-2^64^, hepatitis C virus^65^, and HIV^66^; disease status^67^, as well as novel genome-to-genome study approaches to unpick the impact of complex host-microbe genetic interactions in exerting selective pressures on viral diversity within hosts^68^.

In conclusion, our study provides a high-resolution framework for investigating the viral evolutionary and clinical consequences in the context of vaccination. Fine-scale genomic analyses revealed that the re-emergence of pre-existing lineages, cryptically circulating diversity, and reassortment among locally circulating non-G1P[8] strains contributed to the re-emergence of Wa-like G1P[8] strains, rather than simply by the emergence of an entirely novel post-vaccine genotype constellation. Genome-wide analyses also identified evidence of adaptive evolution, including the near fixation of the VP7:N147D mutation at an antigenically important site within the predominant post-vaccine Wa-like G1P[8] lineage, and, when integrated with clinical data, demonstrated lineage-associated differences in gastroenteritis severity. These findings illustrate how conventional genotype surveillance can obscure epidemiologically and clinically relevant viral diversity and emphasise the value of integrating high-resolution genomics with vaccination, epidemiological, and clinical data. Such approaches will be increasingly important for detecting emerging viral populations, identifying potential antigenic adaptation, and determining whether evolutionary changes arising in the context of vaccines and other clinical interventions translate into meaningful changes in disease phenotype.

### Limitations of the study

Our study has several limitations. First, although linking lineage-level genomic variation with clinical severity provides a novel framework for investigating RVA evolution, we lacked detailed clinical severity data for the pre-vaccine period, preventing direct comparison of disease severity before and after vaccine introduction. Second, the relatively limited number of viruses with detailed clinical information restricted our ability to comprehensively evaluate additional viral, host, epidemiological, and environmental determinants of disease severity. Although the association between the post-vaccine G1P[8] lineages and disease severity persisted after adjusting for age, sex, vaccination status, and number of vaccine doses, residual confounding may remain. In particular, genetic differences elsewhere in the viral genome could have contributed to the observed phenotype. Notably, however, the lineage associated with greater disease severity occurred among the more highly vaccinated group, suggesting that lower vaccine uptake is unlikely to fully explain the higher Vesikari scores observed in this group. Differences in the periods during which the two lineages circulated also limit comparisons of clinical disease severity, as temporal changes in viral, host, epidemiological, or environmental factors may have contributed to the observed difference in disease severity. Comparing lineages co-circulating contemporaneously would likely have provided stronger evidence that the observed differences reflected lineage-specific viral characteristics. Third, periods during which individual lineages were not detected cannot definitively establish cryptic circulation, as incomplete sampling or unsampled introductions could produce similar patterns. Finally, while previous studies suggest a role for residue 147 in antibody recognition and VP7:N147D has been detected in the post-vaccine G1P[8] strains elsewhere, functional studies are required to establish its biological significance in the Malawian population. Neutralisation assays using vaccine-induced antibodies, complemented where feasible by gain- and loss-of-function approaches, will be important to determine whether VP7:N147D directly influences antibody escape, viral fitness, and disease phenotype.

## Methods

### Ethics statement

The National Health Sciences Research Committee, Lilongwe, Malawi, granted ethical approval for the study (protocol number: 867). All study activities were conducted in accordance with the Declaration of Helsinki.

### Sample collection and rotavirus strain selection for whole genome sequencing

We collected stool samples from children under five years who presented with acute gastroenteritis through a long-running sentinel hospital-based surveillance which started in 1997 at the QECH, in Blantyre district, Malawi. We defined acute gastroenteritis as passing at least three looser-than-normal stools in a 24-hour period^69^. We confirmed RVA using a Rotaclone® Enzyme Immunosorbent Assay (Rotaclone®, Meridian Bioscience, Cincinnati, OH, USA). We performed dual genotyping of the VP7 and VP4 genomic segments using a multiplex heminested reverse transcriptase–polymerase chain reaction (PCR) as previously described^35,70,71^. In the present study, we sampled at least one G1P[8] strain per month from 2015 to 2019, following the introduction of the Rotarix vaccine in Malawi. We sampled more than one strain per calendar month in the years when many G1P[8] strains circulated. Genotype and whole-genome sequencing data were already available for G1P[8] rotavirus strains circulating in the pre-vaccine period (1998 to 2012) as well as the early post-vaccine period from 2013 to 2014^36^ and part of 2015^72^.

### Extraction of RVA dsRNA, synthesis of complementary DNA, library preparation and whole-genome sequencing

We extracted RVA dsRNA from raw stool samples using the TRIzol protocol as previously described^36,73^. We removed single-stranded RNA using a lithium chloride (Sigma-Aldrich, Dorset, UK) precipitation for 16 hours at 4 degrees Celsius, followed by DNase 1 (Sigma-Aldrich, Dorset, UK) treatment to remove contaminating DNA^36^. We measured nucleic acid concentration on a Qubit 3.0 fluorometer (Life Technologies, CA, USA). We then assessed dsRNA integrity by electrophoresis on a 1 percent, 0.5× Tris-borate-ethylenediaminetetraacetic acid agarose gel (Sigma-Aldrich, Dorset, UK) stained with SYBR green (Sigma-Aldrich, Dorset, UK) and visualised the gel on a BioDoc transilluminator (Analytik Jena, Germany). We used purified dsRNA to synthesise complementary DNA using the Maxima H Minus Double-Stranded cDNA synthesis kit (Thermo Fisher Scientific, Waltham, MA, USA). Next, we used the MSB® Spin PCRapace (Stratec) purification kit to purify the generated cDNA as previously described^74,75^. We then prepared whole-genome sequencing libraries using the Nextera XT DNA Library Preparation Kit (Illumina, San Diego, CA, USA). We used the Nextera® transposase enzyme to target genomic DNA amplified using a limited-cycle PCR. We used AMPure XP magnetic beads (Beckman Coulter, Pasadena, CA, USA) and 80 percent alcohol to clean up the DNA libraries. To quantify the cleaned-up DNA libraries, we used the Qubit 3.0 fluorometer (Life Technologies, Singapore). We subsequently assessed the fragment size and the quality of libraries using Agilent 2100 BioAnalyzer® (Agilent Technologies, Waldbronn, Germany). Lastly, we generated paired-end nucleotide sequences on a MiSeq® sequencer (Illumina, San Diego, CA, USA) at the University of the Free State’s Next Generation Sequencing (UFS-NGS) Unit, Bloemfontein, South Africa, as previously described^74,75^.

### Illumina reads quality control, sequence assembly and whole genome genotype determination

We trimmed off adapter sequences from raw Illumina sequence reads using Cutadapt *v1.18*^76^. Next, we used FastQC *v0.12.1*^77^ to assess sequence read quality. We further trimmed low-quality bases with Trimmomatic *v0.39*^78^, then filtered reads with a Phred score below 30 to ensure high-quality data for downstream analyses. We mapped the high-quality reads to the prototype Wa-like G1P[8] RVA reference genome (GenBank accession numbers: JX406747.1–JX406757.1) using Geneious Read Mapper *v6.0.3*^79^. We mapped the reads with medium sensitivity and iteratively fine-tuned the parameters five times in Geneious Prime software as previously described^70^. We assigned whole-genome genotype constellations to each individual segment using the Rotavirus A Genotyping Tool *v0.1*^80^. We reported strain genotype constellations based on the full genomes using the previously defined standard nomenclature: Gx-P[x]-Ix-Rx-Cx-Mx-Ax-Nx-Tx-Ex-Hx, corresponding to the following individual segments: VP7-VP4-VP6-VP1-VP2-VP3-NSP1-NSP2-NSP3-NSP4-NSP5, respectively^37^.

### Construction of the standardised Malawi-specific lineage nomenclature database to capture local strain dynamics with finer resolution

To resolve genetic diversity below the conventional genotype level, we defined a standardised, segment-specific lineage nomenclature for all available RVA genome sequences from Malawi to capture strain dynamics effectively over two decades of surveillance. We defined lineages for each genome segment as individual segments evolving independently and therefore with distinct evolutionary histories. By defining segment-specific lineages based on RVA sequences from all genotypes (G1P[8] and non-G1P[8] alike), we captured genetic diversity with finer resolution, including potential intra- and inter-genotype reassortment events. To assign the defined local lineages for each segment based on local lineage nomenclature, we used Viral Lineage Classifier (ViralLC) *v1.1.5*^81^. We used a 98.5% nucleotide identity threshold to define local segment-specific lineages. Sequences sharing at least 98.5% nucleotide identity belonged to the same lineage, whereas those below this threshold were considered putative novel lineages warranting further investigation to confirm the lineages. We used a cutoff of 98.5% to effectively capture with finer resolution temporal changes in lineage turnover and possibly importations of novel lineages in a local setting. We named the local lineages using a local nomenclature comprising the country designation, genome segment, and lineage number. These lineage designations describe locally defined segment-specific lineages within Malawi to address unique locally relevant research questions and should not be used to assign RVA lineages globally, as this will be done using a separate standardised global RVA lineage nomenclature. We included sequences collected across the entire surveillance period when defining the local lineages to ensure newly sequenced strains from Malawi could be classified and easily compared to historical and contemporaneous strains.

### Sub-genotypic RVA lineage assignment and phylogenetic validation

We assigned local lineages to each of the 11 genome segments using the defined local segment-specific RVA lineage nomenclature database from Malawi included in ViralLC *v1.1.5*^81^. We then summarised and visualised the distribution of the local lineages using ggplot2 *v4.0.3*^82^ in R *v4.4.1*^83^. To evaluate the phylogenetic validity of the sequence similarity-based assignments, we constructed segment-specific maximum-likelihood phylogenies for Malawian G1P[8] strains collected over two decades of RVA surveillance (1998-2019). Briefly, we aligned full-length nucleotide sequences for each genome segment independently using MUSCLE *v5.1*^84^. We then inferred maximum-likelihood phylogenies using IQ-TREE2 *v2.1.4-beta*^85^. We used ModelFinder Plus to select the best-fitting nucleotide substitution model for each segment based on the Bayesian Information Criterion. We selected K3Pu+F+G4 model for VP7, VP4, VP3, NSP1, NSP2, NSP4, and NSP5; TIM+F+G4 model for VP1 and VP2; TN+F+R2 for VP6; and TIM+F+I model for NSP3. We assessed the branch support using 1,000 ultrafast bootstrap replicates. We then imported the resulting phylogenies into R *v4.4.1*^83^ using treeio *v1.31.0*^86^, rooted them at the midpoint of the branch connecting the most divergent taxa in the phylogeny using phytools *v2.5-2*^87^, ladderised them to order the nodes using APE *v5.8-1*^88^, and generated visualisations using ggtree *v4.2.0*^89^. We validated the lineage designations by checking whether sequences assigned to the same lineage formed distinct, well-supported monophyletic clusters in the phylogeny, consistent with our 98.5% nucleotide identity threshold for defining local lineages.

### Temporal signal evaluation, time-resolved phylogenetic reconstruction, and tMRCA estimation

Next, we evaluated temporal signal independently for each genome segment using only G1P[8] rotaviruses possessing the canonical Wa-like genotype constellation (G1-P[8]-I1-R1-C1-M1-A1-N1-T1-E1-H1) collected in Malawi between 1998 and 2019. For DS-1-like reassortant G1P[8] viruses (G1-P[8]-I2-R2-C2-M2-A2-N2-T2-E2-H2), we included only the VP7 (G1) and VP4 (P[8]) in the analysis, as they matched the G- and P-genotypes of the canonical Wa-like strains. We excluded the remaining DS-1-like segments because their distinct evolutionary histories could distort segment-specific molecular clock and other downstream phylogenetic analyses. Briefly, we aligned segment-specific nucleotide sequences against the corresponding segment of a prototype G1P[8] strain as the reference (GenBank accession numbers JX406747.1–JX406757.1) using Nextalign *v2.14.0*^90^. We restricted the segment-specific alignments to the open reading frame, and we then reconstructed maximum-likelihood phylogenies as described above. We rooted the generated phylogenies using the oldest ancestral strain with TreeTime *v0.11.1*^91^ and imported the rooted trees into R *v4.4.1*^83^ using treeio *v1.31.0*^86^. We calculated the root-to-tip genetic divergence for each sequence in the phylogeny as the total branch length from the root to the terminal tip using APE *v5.8.1*^88^. We regressed root-to-tip divergence against sampling time using linear regression and then assessed temporal signal based on the estimated regression slope (approximating evolutionary rate in substitutions per site per year), 95% confidence interval, coefficient of determination (*R²*), and the corresponding p-value for the regression coefficient. We identified temporal outliers using standardised regression residuals and flagged sequences with an absolute z-score >3. We then refitted the regressions after excluding outlier sequences. We extrapolated the regression line back to the x-intercept value as an approximate estimate of the date of the tree root, i.e., time to the most recent common ancestor (tMRCA). We generated the root-to-tip regression plots using ggplot2 *v4.0.1*^82^.

We inferred independent segment-specific time-resolved phylogenies under a maximum-likelihood molecular-clock model incorporating sequence sampling dates using TreeTime *v0.11.1*^91^. We visualised the resulting JavaScript Object Notation (JSON) files in Auspice (https://auspice.us) to extract the estimated tMRCA of the Wa-like G1P[8] lineages circulating post-vaccine introduction. We then imported the resulting JSON outputs into R *v4.4.1* using treeio *v1.31.0*^86^, time-scaled branch lengths from node dates expressed in years, and then visualised the dated phylogenies using ggtree *v3.14.0*^92^.

### Selection pressure analysis

We extracted the open reading frames (ORFs) of each of the 11 genome segments by aligning ORFs from the Malawian strains to those of the prototype reference sequences (GenBank accession numbers: JX406747.1 to JX406757.1) using Nextalign *v2.14.0*^90^. We used codon-aware sequences to construct independent phylogenies for each genome segment, as described above, and then used them to assess selection pressure for each segment independently. We obtained the overall nonsynonymous-to-synonymous substitution rate ratio (dN/dS, ω) using the Branch-site Unrestricted Statistical Test for Episodic Diversification (BUSTED)^40^ implemented in HyPhy *v1.3.2*^93^. We also used BUSTED to test for evidence of episodic diversifying selection across each genome segment. For genome segments showing evidence of episodic diversifying selection, we assessed site-specific episodic diversifying selection using the Mixed Effects Model of Evolution (MEME)^94^ implemented in HyPhy *v1.3.2*^93^.

### Protein analysis

For VP7, we aligned the ORFs of Wa-like G1P[8] strains translated to amino acid sequences to the oldest Wa-like G1P[8] strain sequenced in Malawi (GenBank accession number: MG181232.1) as well as to the VP7 sequence of the Wa-like G1P[8] strain included in the Rotarix vaccine using MAFFT *v7.526*^95^. We processed all the multiple sequence alignments in R *v4.4.1*^83^ using Biostrings *v2.80.0*^96^ and Tidyverse *v2.0.0*^97^ to extract amino acid variations at sites showing evidence of episodic diversifying selection in the MEME analysis. We summarised each amino acid substitution at MEME sites against the reference sequences for all the principal VP7 lineages characterised in the current study. We then calculated the proportion of strains carrying each substitution relative to the total number of strains within each lineage. Finally, we visualised the distribution and frequency of amino acid substitutions across VP7 lineages using lineage-level heatmaps generated with ggplot2 *v4.0.3*^82^. To further characterise amino acid differences among the principal VP7 lineages, we selected representative sequences from each lineage and visualised amino acid substitutions relative to the Rotarix VP7 sequence using Snipit *v1.2*^98^. We generated amino acid sequence alignments using Snipit *v1.2*^98^ to show lineage-specific differences across VP7, with particular attention to residues identified as undergoing episodic diversifying selection identified by MEME. To assess the impact of amino acid changes on VP7 protein structure, we generated models for lineages circulating in Malawi and the Rotarix vaccine strain using Swiss-Model^99^. We visualised and annotated the protein models using ChimeraX *v1.12rc202606082314*^100,101^.

### Quantification and statistical analysis

We performed statistical analyses using R *v4.4.1*^83^. We quantified disease severity using the Vesikari scoring system, incorporating diarrhoeal duration and frequency, vomiting duration and frequency, temperature, dehydration, and treatment requirements^46^. We classified disease severity according to established Vesikari score thresholds as mild (<7), moderate (7–10), or severe (≥11)^46^. We expressed Vesikari scores for children infected with strains carrying the principal post-vaccine VP7 lineages as medians and interquartile ranges (IQRs) and compared them using the Wilcoxon rank-sum test. We further assessed the association between VP7 lineage and Vesikari score using multivariable linear regression, adjusting for age, sex, and rotavirus vaccination status. We compared individual components of the Vesikari score between the VP7 lineages to determine which clinical features contributed to the differences in overall disease severity. We adjusted P-values from these component-level comparisons for multiple testing using the Benjamini–Hochberg false discovery rate (FDR) procedure. We defined statistical significance as p < 0.05.

## Data Availability

All complete sequences generated in this study have been deposited in GenBank under the following accession numbers PX492636 to PX493357 and from PX495251 to PX495327.

## Data availability

We deposited all complete sequences generated in this study in GenBank^102^ (accession numbers: PX492636 to PX493357 and PX495251 to PX495327). This study also used metadata and genome sequences generated in our earlier study (accession numbers: MG181227 to MG181941).

## Supplemental information

- **Figure S1.** Classification and temporal distribution of local G1P[8] non-structural protein (NSP) gene lineages in Malawi, 1998–2019.
- **Figure S2.** Root-to-tip regression analysis of rotavirus genome segments.
- **Figure S3.** Time-resolved phylogenetic reconstruction of G1P[8] rotavirus genome-segment lineages circulating in Malawi.
- **Figure S4.** Phylogeographical analysis of the eleven segments of Malawian and contemporary G1P[8] strains detected elsewhere.
- **Figure S5.** Temporal frequencies of G1P[8] viruses carrying asparagine (N147) or aspartic acid (D147) at VP7 residue 147 in sampled strains by country globally.
- **Figure S6.** Comparison of individual clinical severity components between post-vaccine MW–VP7–L1 and MW–VP7–L2 G1P[8] rotavirus infections.
- **Table S1.** Complete genotype constellation and collection dates for Malawian G1P[8] strains sequenced from 2015-2019.

● **Table S2.** Genomic lineage diversity of G1P[8] rotaviruses in Malawi across the study period and after 2014

## Acknowledgements

We thank the laboratory and clinical research teams within the Global Health and Enteric Disease Research Group at the Malawi-Liverpool-Wellcome Research Programme, as well as the study participants, for their invaluable contributions. We also thank the Centre for Genomic Research at the University of Liverpool and the University of the Free State Next Generation Sequencing Unit (UFS-NGS) for generating genomic data for this project. We further acknowledge the African Enterics Viral Genome Initiative (AEVGI) for facilitating the initial phase of this large-scale rotavirus whole-genome sequencing collaboration among Malawi, Ghana, and South Africa. The Sapitwa High Performance Computing (HPC) Cluster at the Malawi Liverpool Wellcome Research Programme (MLW), funded through the Wellcome core award to MLW (grant number: 206545/Z/17/Z), provided computational resources.

## Author contribution

C.M. and K.C.J. conceived and designed the study. C.C., C.M.D., A.W.K. and K.C.J. supervised the study. K.C.J. secured funds for the study. N.A.C. and K.C.J. collected clinical data and stool samples. C.M., E.C., A.W.K., M.M.N., and K.C.J. performed the laboratory work. M.M.N. and K.C.J. generated the whole-genome sequence data. M.M. provided high-performance computing (HPC) support. R.W. and C.M. curated clinical and laboratory datasets. C.M. designed the analytical plan, provided the software, and carried out the bioinformatic and statistical analyses, with input from C.C. and K.C.J. C.M. wrote the initial draft with input from C.C. and K.C.J. All authors reviewed and approved the manuscript.

