## Supplemental figures for "Genomic epidemiology reveals post-vaccine re-emergence of vaccine-associated G1P[8] rotavirus lineages with variable disease severity"

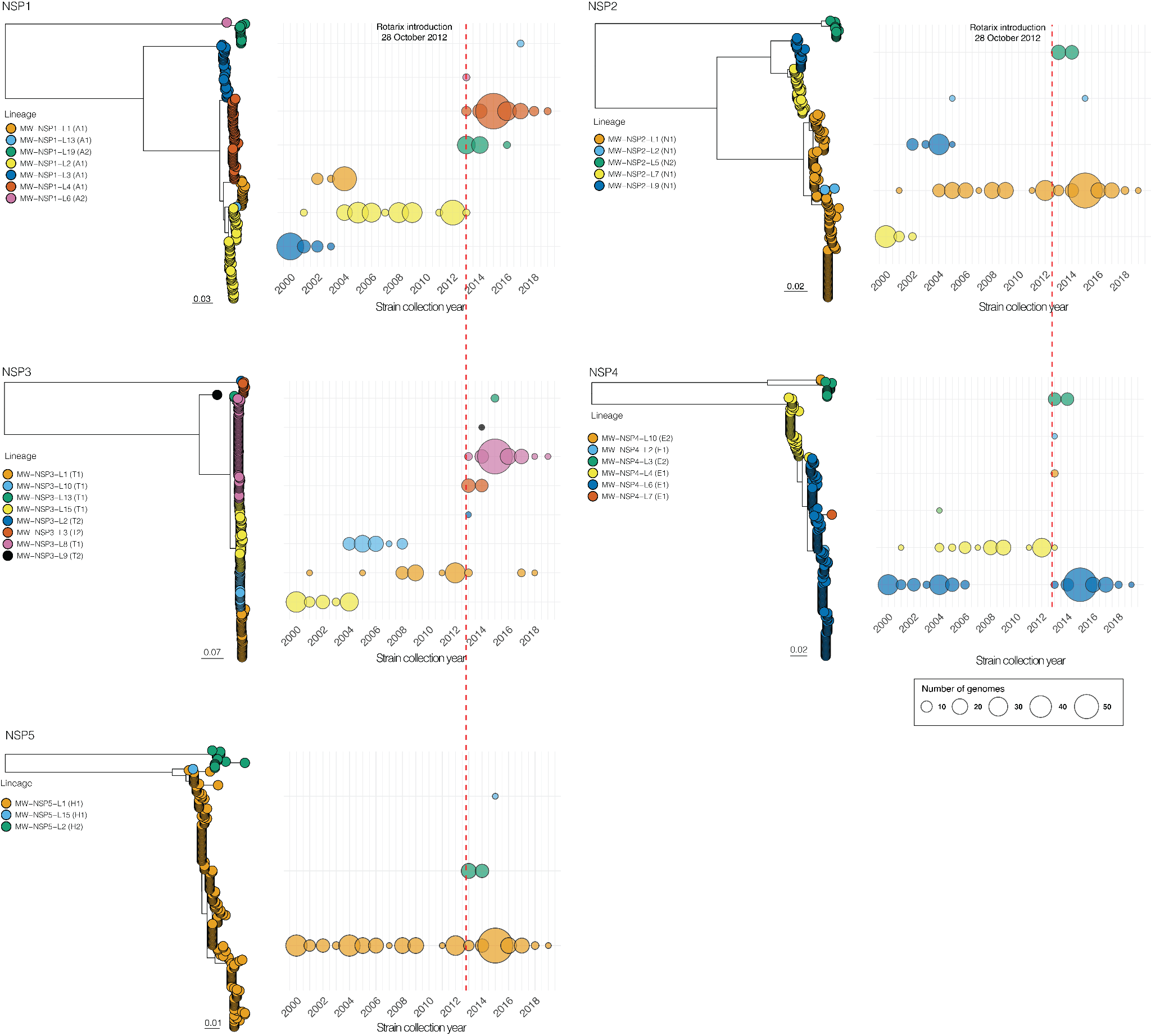

**Figure S1. Classification and temporal distribution of local G1P[8] non-structural protein (NSP) gene lineages in Malawi, 1998–2019.** Maximum-likelihood phylogenies are shown for NSP1, NSP2, NSP3, NSP4 and NSP5 encoding genome segments. Local Malawian lineages were assigned using ViralLC based on nucleotide sequence similarity across viruses sampled between 1998 and 2019; these designations represent locally defined lineages rather than global lineage nomenclature. Phylogenetic tips are coloured according to the assigned local lineage, with the corresponding conventional segment genotype shown in parentheses. For each non-structural protein gene, sequences assigned to the same local lineage are generally clustered together in the maximum-likelihood tree, demonstrating concordance between the sequence-similarity-based classification and the inferred phylogenetic structure. The adjacent bubble plots show the annual distribution of each lineage, with bubble size proportional to the number of genomes detected in each surveillance year. The vertical dashed red line marks the introduction of Rotarix rotavirus vaccine into the national immunisation program in Malawi on 28 October 2012. Phylogenetic scale bars indicate nucleotide substitutions per site

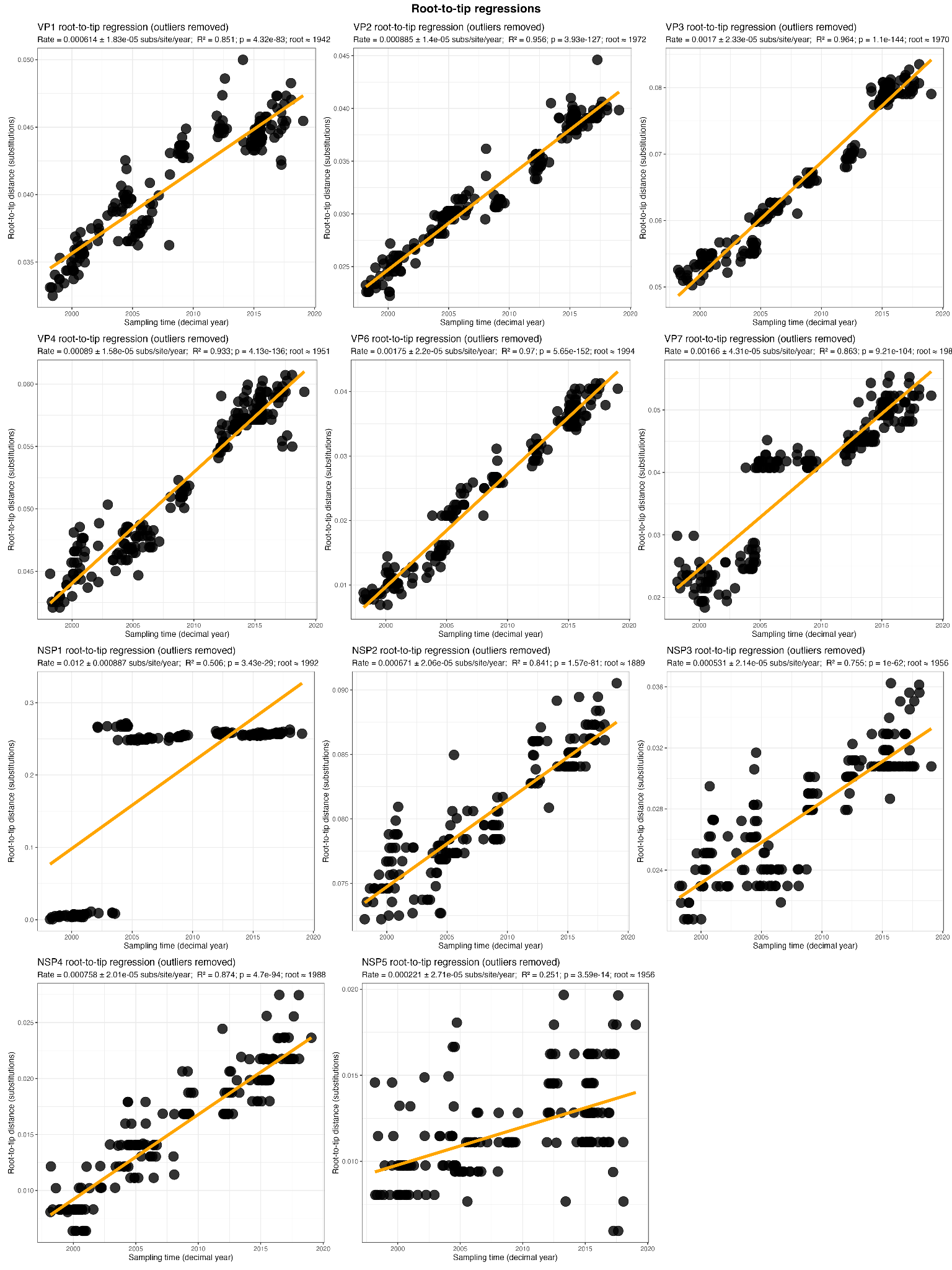

**Figure S2. Root-to-tip regression analysis of rotavirus genome segments.** Root-to-tip genetic distances were regressed against sampling time for each of the 11 rotavirus genome segments (VP1–VP4, VP6, VP7, and NSP1–NSP5) after removal of identified temporal outliers. Each point represents an individual sequence, and the fitted line represents the linear relationship between sampling date and root-to-tip divergence. The slope of each regression provides an approximate evolutionary rate in substitutions per site per year, while R^2 and the corresponding *p*-value quantify the strength and statistical significance of the temporal signal.

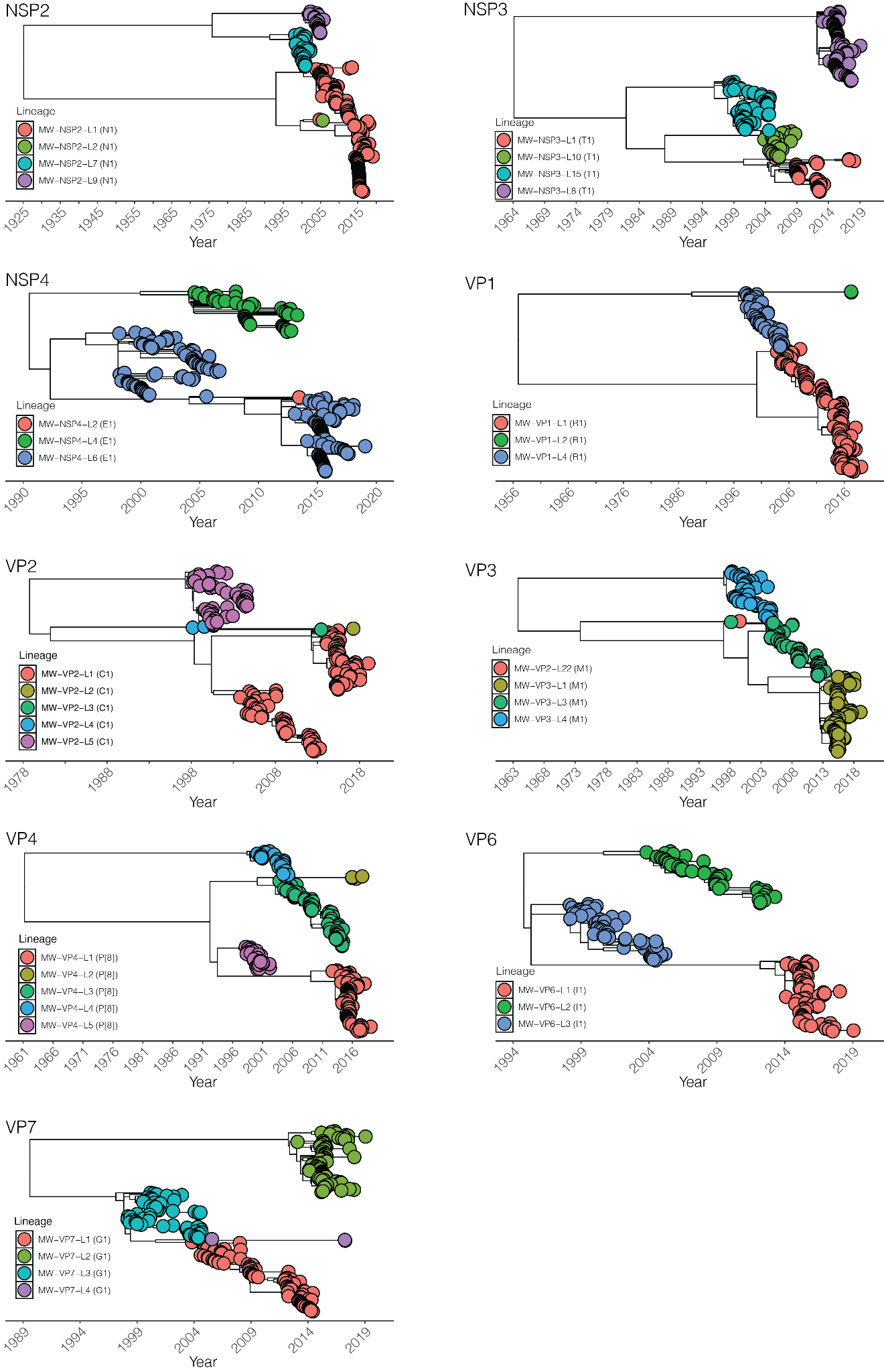

**Figure S3. Time-resolved phylogenetic reconstruction of G1P[8] rotavirus genome-segment lineages circulating in Malawi.** Time-scaled phylogenetic trees are shown for NSP2, NSP3, NSP4, VP1, VP2, VP3, VP4, VP6, and VP7 encoding genome segments. Branch lengths are scaled according to calendar time, with the x-axis indicating year of inferred ancestry or sampling. Tips are coloured according to the corresponding Malawi segment-specific lineage, as indicated in the legend of each panel. The phylogenies illustrate the distinct evolutionary histories of individual genome segments and show that lineages detected during the post-vaccine period arose at different times, with several having ancestral origins predating their observed circulation. The coexistence of segment lineages with differing temporal histories is consistent with the assembly of G1P[8] genomes from independently evolving segment lineages.

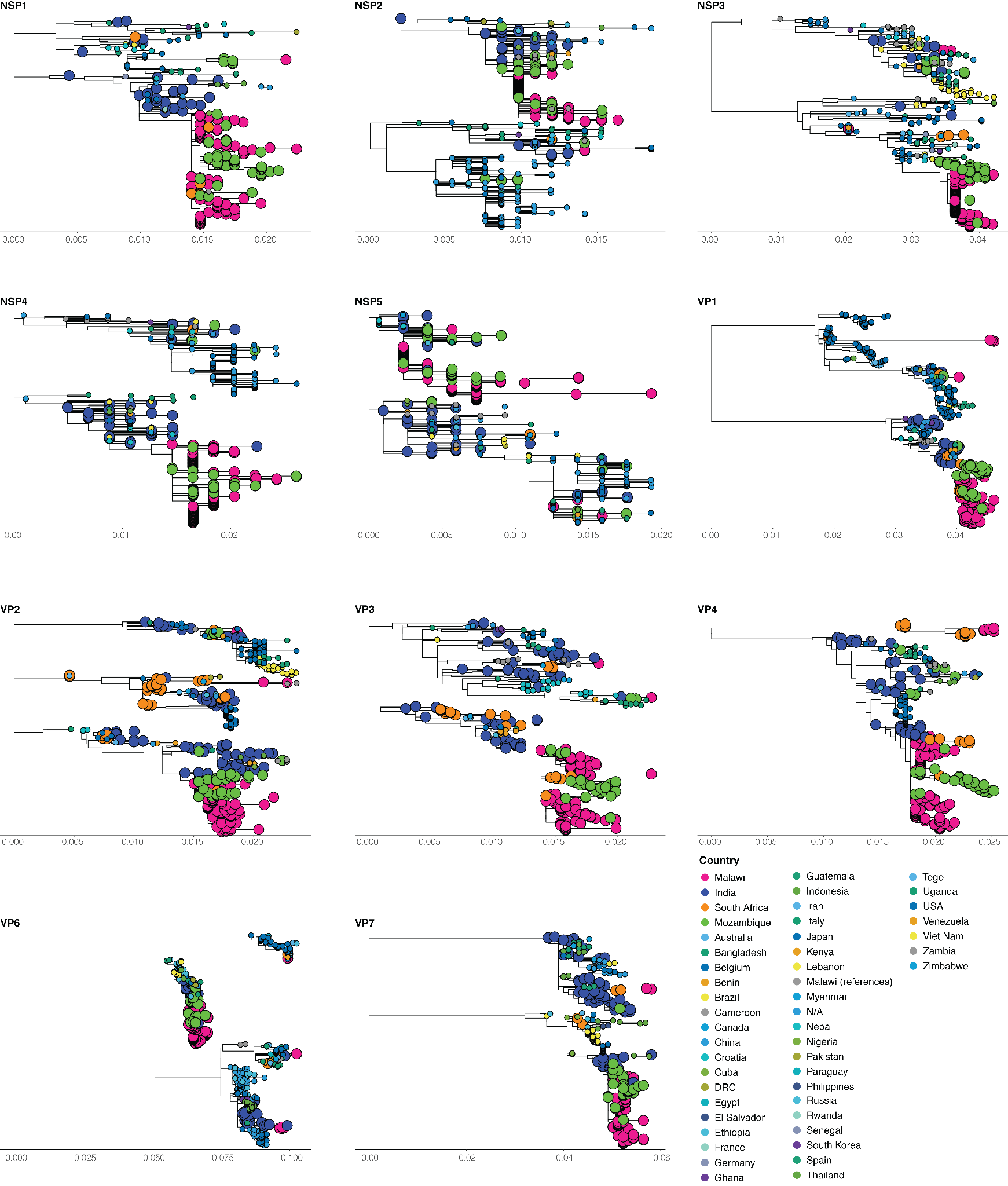

**Figure S4. Phylogeographical analysis of the eleven segments of Malawian and contemporary G1P[8] strains detected elsewhere**. Analyses were restricted to sequences with complete open reading frames. Tip labels are annotated by country of sampling. Malawian strains generated in this study and strains from countries clustering together with Malawian strains are highlighted with bigger tip nodes. Trees were midpoint-rooted, and node support values represent ultrafast bootstrap support from 1,000 replicates.

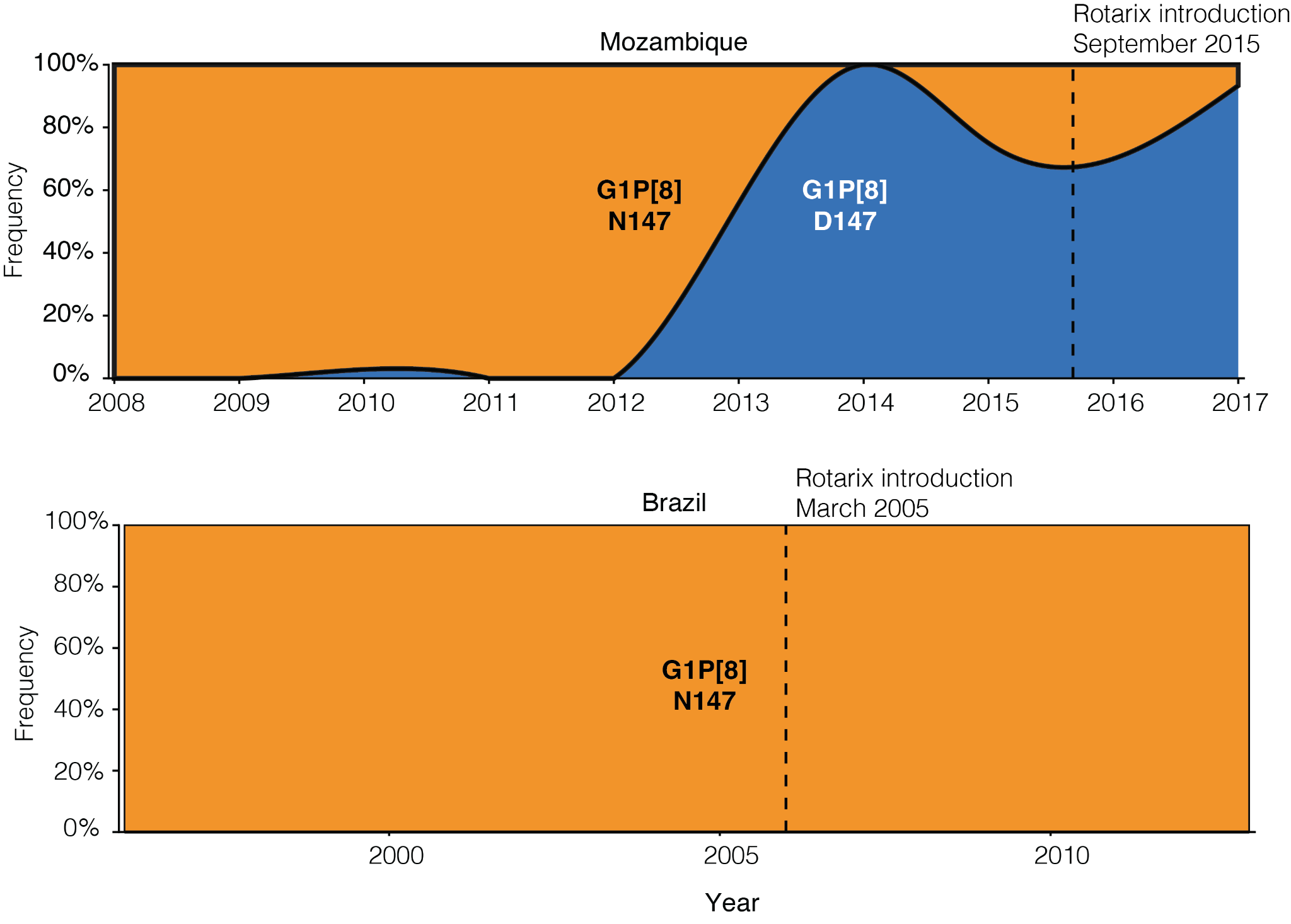
**Figure S5. Temporal frequencies of G1P[8] viruses carrying asparagine (N147) or aspartic acid (D147) at VP7 residue 147 in sampled Rotarix using countries across the globe.** Residue frequencies were calculated from available G1P[8] VP7 sequences by year of isolation, with vertical dashed lines indicating Rotarix vaccine introduction in each setting.

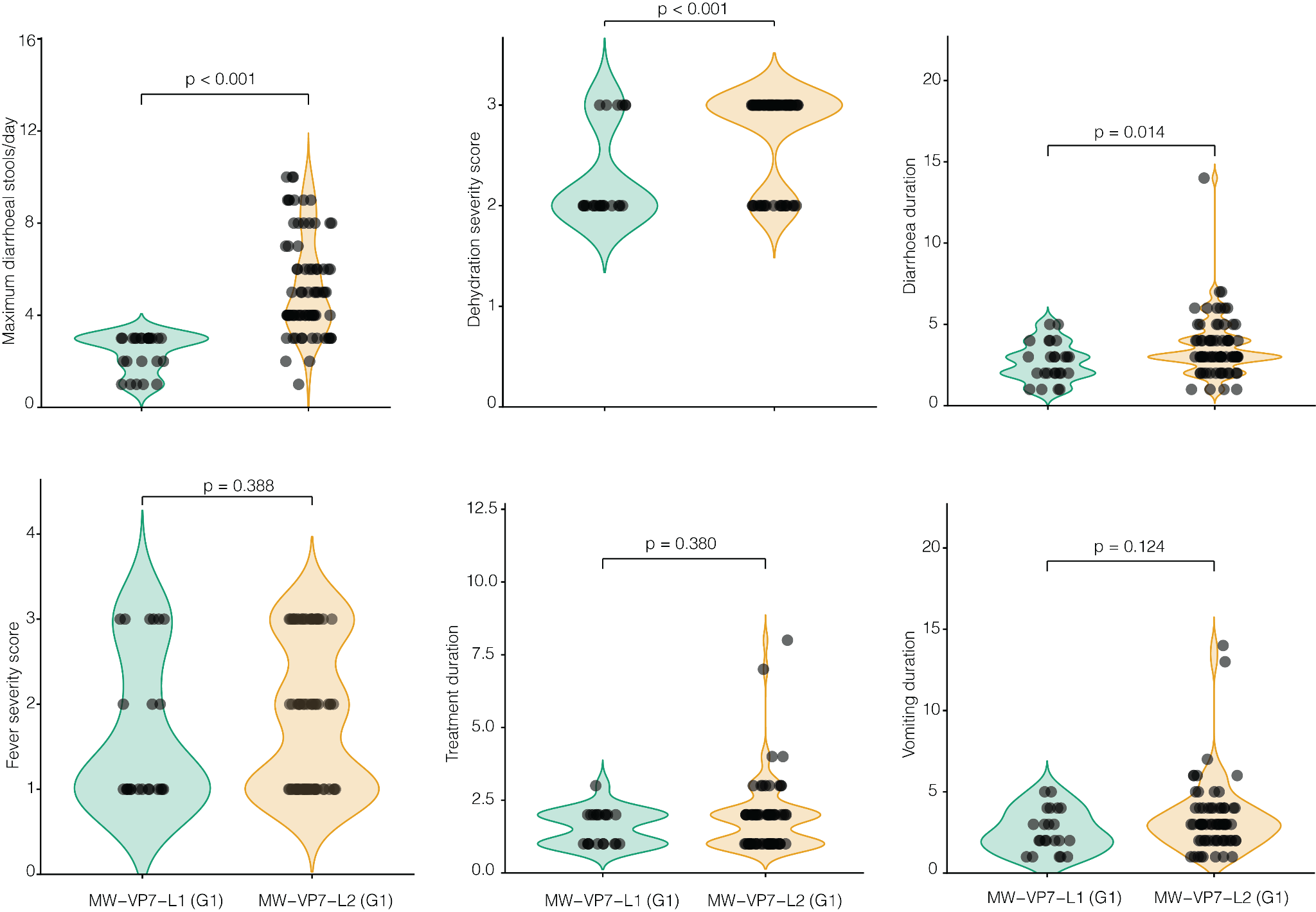

**Figure S6. Comparison of individual clinical severity components between post-vaccine MW–VP7–L1 and MW–VP7–L2 G1P[8] rotavirus infections.** Violin plots show the distributions of six components contributing to clinical disease severity among children infected with MW–VP7–L1 (G1) and MW–VP7–L2 (G1) strains.
