## Supplemental tables for "Genomic epidemiology reveals post-vaccine re-emergence of vaccine-associated G1P[8] rotavirus lineages with variable disease severity"

**Table S1 Complete genotype constellation and collection dates for Malawian G1P[8] strains sequenced from 2015-2019.**

| Strain | Collection  date | VP7 | VP4 | VP6 | VP1 | VP2 | VP3 | NSP1 | NSP2 | NSP3 | NSP4 | NSP5/6 |
| --- | --- | --- | --- | --- | --- | --- | --- | --- | --- | --- | --- | --- |
| RVA/Human-wt/MWI/BTY104/2015/G1P[8] | 26-Jan-15 | G1 | P[8] | I1 | R1 | - | M1 | - | N1 | T1 | E1 | H1 |
| RVA/Human-wt/MWI/BTY103/2015/G1P[8] | 26-Jan-15 | G1 | P[8] | I1 | R1 | C1 | M1 | - | N1 | T1 | E1 | H1 |
| RVA/Human-wt/MWI/BTY10A/2015/G1P[8] | 27-Jan-15 | G1 | P[8] | I1 | - | - | M1 | A1 | N1 | T1 | E1 | H1 |
| RVA/Human-wt/MWI/BTY105/2015/G1P[8] | 30-Jan-15 | G1 | P[8] | I1 | R1 | C1 | M1 | A1 | N1 | T1 | E1 | H1 |
| RVA/Human-wt/MWI/BTY10G/2015/G1P[8] | 30-Jan-15 | G1 | P[8] | I1 | R1 | C1 | M1 | - | N1 | T1 | E1 | H1 |
| RVA/Human-wt/MWI/BTY10W/2015/G1P[8] | 03-Feb-15 | G1 | P[8] | I1 | R1 | - | M1 | - | N1 | T1 | E1 | H1 |
| RVA/Human-wt/MWI/BTY10T/2015/G1P[8] | 03-Feb-15 | G1 | P[8] | I1 | R1 | C1 | M1 | A1 | N1 | T1 | E1 | H1 |
| RVA/Human-wt/MWI/BTY10S/2015/G1P[8] | 03-Feb-15 | G1 | P[8] | I1 | R1 | C1 | M1 | A1 | N1 | T1 | E1 | H1 |
| RVA/Human-wt/MWI/BTY10Y/2015/G1P[8] | 04-Feb-15 | G1 | P[8] | I1 | R1 | - | M1 | - | N1 | T1 | E1 | - |
| RVA/Human-wt/MWI/BTY12V/2015/G1P[8] | 04-Mar-15 | G1 | P[8] | I1 | R1 | C1 | M1 | A1 | N1 | T1 | E1 | H1 |
| RVA/Human-wt/MWI/BTY135/2015/G1P[8] | 09-Mar-15 | G1 | P[8] | I1 | R1 | C1 | M1 | A1 | N1 | T1 | E1 | H1 |
| RVA/Human-wt/MWI/BTY13C/2015/G1P[8] | 10-Mar-15 | G1 | P[8] | I1 | R1 | C1 | M1 | A1 | N1 | T1 | E1 | H1 |
| RVA/Human-wt/MWI/BTY13P/2015/G1P[8] | 17-Mar-15 | G1 | P[8] | I1 | R1 | C1 | M1 | A1 | N1 | T1 | E1 | H1 |
| RVA/Human-wt/MWI/BTY13W/2015/G1P[8] | 19-Mar-15 | G1 | P[8] | I1 | R1 | C1 | M1 | A1 | N1 | T1 | E1 | H1 |
| RVA/Human-wt/MWI/BTY142/2015/G1P[8] | 23-Mar-15 | G1 | P[8] | I1 | R1 | C1 | M1 | A1 | N1 | T1 | E1 | H1 |
| RVA/Human-wt/MWI/BTY14N/2015/G1P[8] | 30-Mar-15 | G1 | P[8] | I1 | R1 | C1 | M1 | A1 | N1 | T1 | E1 | H1 |
| RVA/Human-wt/MWI/BTY18G/2015/G1P[8] | 07-Apr-15 | G1 | P[8] | I1 | R1 | C1 | M1 | A1 | N1 | T1 | E1 | H1 |
| RVA/Human-wt/MWI/BTY18K/2015/G1P[8] | 08-Apr-15 | G1 | P[8] | I1 | R1 | C1 | M1 | A1 | N1 | T1 | E1 | H1 |
| RVA/Human-wt/MWI/BTY18Q/2015/G1P[8] | 14-Apr-15 | G1 | P[8] | I1 | R1 | C1 | M1 | A1 | N1 | T1 | E1 | H1 |
| RVA/Human-wt/MWI/BTY19A/2015/G1P[8] | 22-Apr-15 | G1 | P[8] | I1 | R1 | C1 | M1 | A1 | N1 | T1 | E1 | H1 |
| RVA/Human-wt/MWI/BTY19H/2015/G1P[8] | 28-Apr-15 | G1 | P[8] | I1 | R1 | - | M1 | A1 | N1 | - | E1 | H1 |
| RVA/Human-wt/MWI/BTY1A3/2015/G1P[8] | 12-May-15 | G1 | P[8] | I1 | R1 | C1 | M1 | - | N1 | T1 | E1 | H1 |
| RVA/Human-wt/MWI/BTY1A7/2015/G1P[8] | 13-May-15 | G1 | P[8] | I1 | R1 | C1 | M1 | A1 | N1 | T1 | E1 | H1 |
| RVA/Human-wt/MWI/BTY1A9/2015/G1P[8] | 15-May-15 | G1 | P[8] | I1 | R1 | - | M1 | A1 | N1 | T1 | E1 | H1 |
| RVA/Human-wt/MWI/BTY1AC/2015/G1P[8] | 18-May-15 | G1 | P[8] | I1 | R1 | - | - | A1 | N1 | - | E1 | H1 |
| RVA/Human-wt/MWI/BTY1AJ/2015/G1P[8] | 20-May-15 | G1 | P[8] | I1 | R1 | C1 | M1 | - | N1 | T1 | E1 | H1 |
| RVA/Human-wt/MWI/BTY1AK/2015/G1P[8] | 21-May-15 | G1 | P[8] | I1 | R1 | C1 | M1 | - | N1 | T1 | E1 | H1 |
| RVA/Human-wt/MWI/BTY1AV/2015/G1P[8] | 28-May-15 | G1 | P[8] | I1 | R1 | C1 | M1 | A1 | N1 | T1 | E1 | H1 |
| RVA/Human-wt/MWI/BTY1B2/2015/G1P[8] | 02-Jun-15 | G1 | P[8] | I1 | R1 | - | M1 | A1 | N1 | T1 | E1 | H1 |
| RVA/Human-wt/MWI/BTY1B4/2015/G1P[8] | 04-Jun-15 | G1 | P[8] | I1 | R1 | C1 | M1 | - | N1 | T1 | E1 | H1 |
| RVA/Human-wt/MWI/BTY1BR/2015/G1P[8] | 16-Jun-15 | G1 | P[8] | I1 | R1 | C1 | M1 | - | N1 | T1 | E1 | H1 |
| RVA/Human-wt/MWI/BTY1BS/2015/G1P[8] | 17-Jun-15 | G1 | P[8] | I1 | R1 | C1 | M1 | - | N1 | T1 | E1 | H1 |
| RVA/Human-wt/MWI/BTY1C1/2015/G1P[8] | 19-Jun-15 | G1 | P[8] | I1 | R1 | C1 | M1 | - | N1 | T1 | E1 | H1 |
| RVA/Human-wt/MWI/BTY1BY/2015/G1P[8] | 23-Jun-15 | G1 | P[8] | I1 | R1 | - | M1 | - | N1 | T1 | E1 | H1 |
| RVA/Human-wt/MWI/BTY1BX/2015/G1P[8] | 23-Jun-15 | G1 | P[8] | I1 | R1 | C1 | M1 | A1 | N1 | T1 | E1 | H1 |
| RVA/Human-wt/MWI/BTY1CT/2015/G1P[8] | 01-Jul-15 | G1 | P[8] | I1 | R1 | - | M1 | - | N1 | T1 | E1 | H1 |
| RVA/Human-wt/MWI/BTY1CP/2015/G1P[8] | 01-Jul-15 | G1 | P[8] | I1 | R1 | C1 | M1 | A1 | N1 | T1 | E1 | H1 |
| RVA/Human-wt/MWI/BTY1CW/2015/G1P[8] | 02-Jul-15 | G1 | P[8] | I1 | R1 | - | - | - | N1 | T1 | E1 | H1 |
| RVA/Human-wt/MWI/BTY1D1/2015/G1P[8] | 07-Jul-15 | G1 | P[8] | I1 | R1 | - | M1 | A1 | N1 | T1 | E1 | H1 |
| RVA/Human-wt/MWI/BTY1D6/2015/G1P[8] | 13-Jul-15 | G1 | P[8] | I1 | R1 | C1 | M1 | A1 | N1 | T1 | E1 | H1 |
| RVA/Human-wt/MWI/BTY14W/2015/G1P[8] | 14-Jul-15 | G1 | P[8] | I1 | R1 | - | M1 | - | N1 | T1 | E1 | H1 |
| RVA/Human-wt/MWI/BTY152/2015/G1P[8] | 15-Jul-15 | G1 | P[8] | I1 | R1 | C1 | M1 | A1 | N1 | T1 | E1 | H1 |
| RVA/Human-wt/MWI/BTY158/2015/G1P[8] | 20-Jul-15 | G1 | P[8] | I1 | R1 | C1 | M1 | A1 | N1 | T1 | E1 | H1 |
| RVA/Human-wt/MWI/BTY15D/2015/G1P[8] | 24-Jul-15 | G1 | P[8] | I1 | R1 | C1 | M1 | A1 | N1 | T1 | - | H1 |
| RVA/Human-wt/MWI/BTY15E/2015/G1P[8] | 27-Jul-15 | G1 | P[8] | I1 | R1 | C1 | M1 | A1 | N1 | T1 | E1 | H1 |
| RVA/Human-wt/MWI/BTY15Y/2015/G1P[8] | 10-Aug-15 | G1 | P[8] | I1 | R1 | - | M1 | A1 | N1 | T1 | E1 | H1 |
| RVA/Human-wt/MWI/BTY163/2015/G1P[8] | 12-Aug-15 | G1 | - | I1 | R1 | - | M1 | A1 | N1 | T1 | E1 | H1 |
| RVA/Human-wt/MWI/BTY16E/2015/G1P[8] | 19-Aug-15 | G1 | P[8] | I1 | R1 | - | M1 | A1 | N1 | T1 | E1 | H1 |
| RVA/Human-wt/MWI/BTY16L/2015/G1P[8] | 26-Aug-15 | G1 | P[8] | I1 | R1 | C1 | M1 | A1 | N1 | T1 | E1 | H1 |
| RVA/Human-wt/MWI/BTY16X/2015/G1P[8] | 04-Sep-15 | G1 | P[8] | I1 | R1 | - | M1 | A1 | N1 | - | E1 | H1 |
| RVA/Human-wt/MWI/BTY16W/2015/G1P[8] | 04-Sep-15 | G1 | P[8] | I1 | R1 | - | M1 | A1 | N1 | T1 | E1 | H1 |
| RVA/Human-wt/MWI/BTY174/2015/G1P[8] | 08-Sep-15 | G1 | P[8] | I1 | R1 | - | M1 | A1 | - | T1 | E1 | H1 |
| RVA/Human-wt/MWI/BTY17I/2015/G1P[8] | 17-Sep-15 | G1 | P[8] | I1 | R1 | C1 | M1 | A1 | N1 | T1 | E1 | H1 |
| RVA/Human-wt/MWI/BTY1E5/2015/G1P[8] | 09-Dec-15 | G1 | P[8] | I1 | R1 | C1 | M1 | A1 | N1 | T1 | E1 | H1 |
| RVA/Human-wt/MWI/BTY1EQ/2016/G1P[8] | 05-Jan-16 | G1 | P[8] | I1 | R1 | C1 | M1 | A1 | - | T1 | E1 | H1 |
| RVA/Human-wt/MWI/BTY1FF/2016/G1P[8] | 21-Jan-16 | G1 | P[8] | I1 | R1 | C1 | M1 | A1 | N1 | T1 | E1 | H1 |
| RVA/Human-wt/MWI/BTY1L8/2016/G1P[8] | 10-May-16 | G1 | P[8] | I1 | R1 | C1 | M1 | A1 | N1 | T1 | E1 | H1 |
| RVA/Human-wt/MWI/BTY1LV/2016/G1P[8] | 20-Jun-16 | G1 | P[8] | I1 | R1 | C1 | M1 | A1 | N1 | T1 | E1 | H1 |
| RVA/Human-wt/MWI/BTY1M4/2016/G1P[8] | 27-Jun-16 | G1 | P[8] | I1 | R1 | C1 | M1 | A1 | N2 | T1 | E1 | H1 |
| RVA/Human-wt/MWI/BTY1ND/2016/G1P[8] | 08-Sep-16 | G1 | P[8] | I1 | R1 | C1 | M1 | A1 | N1 | T1 | E1 | H1 |
| RVA/Human-wt/MWI/BTY1QX/2016/G1P[8] | 04-Nov-16 | G1 | P[8] | I1 | - | C1 | M1 | A1 | N1 | T1 | E1 | H1 |
| RVA/Human-wt/MWI/BTY1QY/2016/G1P[8] | 08-Nov-16 | G1 | P[8] | I1 | R1 | C1 | M1 | A1 | N1 | T1 | E1 | H1 |
| RVA/Human-wt/MWI/BTY1RW/2016/G1P[8] | 06-Dec-16 | G1 | P[8] | I1 | R1 | C1 | M1 | A1 | N1 | T1 | E1 | H1 |
| RVA/Human-wt/MWI/BTY1RV/2016/G1P[8] | 06-Dec-16 | G1 | P[8] | I1 | R1 | C1 | M1 | A1 | N1 | T1 | E1 | H1 |
| RVA/Human-wt/MWI/BTY1SA/2017/G1P[8] | 04-Jan-17 | G1 | P[8] | I1 | R1 | C1 | M1 | A1 | N1 | T1 | E1 | H1 |
| RVA/Human-wt/MWI/BTY1VH/2017/G1P[8] | 27-Mar-17 | G1 | P[8] | I1 | R1 | C1 | M1 | A1 | N1 | T1 | E1 | H1 |
| RVA/Human-wt/MWI/BTY1VK/2017/G1P[8] | 29-Mar-17 | G1 | P[8] | I1 | R1 | C1 | M1 | A1 | N1 | T1 | E1 | H1 |
| RVA/Human-wt/MWI/BTY1VQ/2017/G1P[8] | 04-Apr-17 | G1 | P[8] | I1 | R1 | C1 | M1 | A1 | N1 | T1 | E1 | H1 |
| RVA/Human-wt/MWI/BTY1VV/2017/G1P[8] | 07-Apr-17 | G1 | P[8] | I1 | R1 | C1 | M1 | A1 | N1 | T1 | E1 | H1 |
| RVA/Human-wt/MWI/BTY1WM/2017/G1P[8] | 08-May-17 | G1 | P[8] | I1 | R1 | C1 | M1 | A1 | N1 | T1 | E1 | H1 |
| RVA/Human-wt/MWI/BTY1YW/2017/G1P[8] | 03-Jul-17 | G1 | P[8] | I1 | R1 | C1 | M1 | A1 | N1 | T1 | E1 | H1 |
| RVA/Human-wt/MWI/BTY1Z6/2017/G1P[8] | 11-Jul-17 | G1 | P[8] | I1 | R1 | C1 | M1 | - | - | T1 | E1 | - |
| RVA/Human-wt/MWI/BTY20Q/2017/G1P[8] | 24-Aug-17 | G1 | P[8] | I1 | R1 | C1 | M1 | - | N1 | T1 | E1 | H1 |
| RVA/Human-wt/MWI/BTY20R/2017/G1P[8] | 25-Aug-17 | G1 | P[8] | I1 | R1 | C1 | M1 | - | N1 | T1 | E1 | H1 |
| RVA/Human-wt/MWI/BTY23Y/2018/G1P[8] | 18-Jan-18 | G1 | P[8] | I1 | R1 | C1 | M1 | A1 | N1 | T1 | E1 | H1 |
| RVA/Human-wt/MWI/BTY246/2018/G1P[8] | 25-Jan-18 | G1 | P[8] | I1 | R1 | C1 | M1 | A1 | N1 | T1 | E1 | H1 |
| RVA/Human-wt/MWI/BTY2B0/2019/G1P[8] | 21-Jan-19 | G1 | P[8] | I1 | R1 | C1 | M1 | A1 | N1 | T1 | E1 | H1 |

**Table S2 Genomic lineage diversity of G1P[8] rotaviruses in Malawi across the study period and after 2014.** For each of the 11 rotavirus genome segments, the table shows the total number of distinct phylogenetically defined Malawian lineages identified across the complete study period and the corresponding lineage designations. Post-2014 summarises the number of these lineages detected among G1P[8] strains circulating from 2015 to 2019 and lists the lineages detected.

| Segment | Overall_n | Overall_lineages | Post_2014_n | Post_2014_lineages |
| --- | --- | --- | --- | --- |
| NSP1 | 7 | MW-NSP1-L1 (A1) | 3 | MW-NSP1-L4 (A1) |
|  |  | MW-NSP1-L2 (A1) |  | MW-NSP1-L13 (A1) |
|  |  | MW-NSP1-L3 (A1) |  | MW-NSP1-L19 (A2) |
|  |  | MW-NSP1-L4 (A1) |  |  |
|  |  | MW-NSP1-L6 (A2) |  |  |
|  |  | MW-NSP1-L13 (A1) |  |  |
|  |  | MW-NSP1-L19 (A2) |  |  |
| NSP2 | 5 | MW-NSP2-L1 (N1) | 2 | MW-NSP2-L1 (N1) |
|  |  | MW-NSP2-L2 (N1) |  | MW-NSP2-L2 (N1) |
|  |  | MW-NSP2-L5 (N2) |  |  |
|  |  | MW-NSP2-L7 (N1) |  |  |
|  |  | MW-NSP2-L9 (N1) |  |  |
| NSP3 | 8 | MW-NSP3-L1 (T1) | 2 | MW-NSP3-L1 (T1) |
|  |  | MW-NSP3-L2 (T2) |  | MW-NSP3-L8 (T1) |
|  |  | MW-NSP3-L3 (T2) |  |  |
|  |  | MW-NSP3-L8 (T1) |  |  |
|  |  | MW-NSP3-L9 (T2) |  |  |
|  |  | MW-NSP3-L10 (T1) |  |  |
|  |  | MW-NSP3-L13 (T1) |  |  |
|  |  | MW-NSP3-L15 (T1) |  |  |
| NSP4 | 6 | MW-NSP4-L2 (E1) | 1 | MW-NSP4-L6 (E1) |
|  |  | MW-NSP4-L3 (E2) |  |  |
|  |  | MW-NSP4-L4 (E1) |  |  |
|  |  | MW-NSP4-L6 (E1) |  |  |
|  |  | MW-NSP4-L7 (E1) |  |  |
|  |  | MW-NSP4-L10 (E2) |  |  |
| NSP5 | 3 | MW-NSP5-L1 (H1) | 2 | MW-NSP5-L1 (H1) |
|  |  | MW-NSP5-L2 (H2) |  | MW-NSP5-L15 (H1) |
|  |  | MW-NSP5-L15 (H1) |  |  |
| VP1 | 5 | MW-VP1-L1 (R1) | 2 | MW-VP1-L1 (R1) |
|  |  | MW-VP1-L2 (R1) |  | MW-VP1-L2 (R1) |
|  |  | MW-VP1-L3 (R1) |  |  |
|  |  | MW-VP1-L4 (R1) |  |  |
|  |  | MW-VP1-L6 (R2) |  |  |
| VP2 | 7 | MW-VP2-L1 (C1) | 2 | MW-VP2-L1 (C1) |
|  |  | MW-VP2-L2 (C1) |  | MW-VP2-L2 (C1) |
|  |  | MW-VP2-L3 (C1) |  |  |
|  |  | MW-VP2-L4 (C1) |  |  |
|  |  | MW-VP2-L5 (C1) |  |  |
|  |  | MW-VP2-L8 (C2) |  |  |
|  |  | MW-VP2-L24 (C1) |  |  |
| VP3 | 6 | MW-VP3-L1 (M1) | 2 | MW-VP3-L1 (M1) |
|  |  | MW-VP3-L2 (M1) |  | MW-VP3-L2 (M1) |
|  |  | MW-VP3-L3 (M1) |  |  |
|  |  | MW-VP3-L4 (M1) |  |  |
|  |  | MW-VP3-L7 (M2) |  |  |
|  |  | MW-VP3-L8 (M2) |  |  |
| VP4 | 5 | MW-VP4-L1 (P[8]) | 2 | MW-VP4-L1 (P[8]) |
|  |  | MW-VP4-L2 (P[8]) |  | MW-VP4-L2 (P[8]) |
|  |  | MW-VP4-L3 (P[8]) |  |  |
|  |  | MW-VP4-L4 (P[8]) |  |  |
|  |  | MW-VP4-L5 (P[8]) |  |  |
| VP6 | 4 | MW-VP6-L1 (I1) | 1 | MW-VP6-L1 (I1) |
|  |  | MW-VP6-L2 (I1) |  |  |
|  |  | MW-VP6-L3 (I1) |  |  |
|  |  | MW-VP6-L5 (I2) |  |  |
| VP7 | 4 | MW-VP7-L1 (G1) | 2 | MW-VP7-L2 (G1) |
|  |  | MW-VP7-L2 (G1) |  | MW-VP7-L4 (G1) |
|  |  | MW-VP7-L3 (G1) |  |  |
|  |  | MW-VP7-L4 (G1) |  |  |
